# Phenome-derived polygenic scores and social determinants jointly shape context-dependent disease risk

**DOI:** 10.64898/2026.04.16.26351039

**Authors:** Ying Wang, Buu Truong, Wenhan Lu, Chaimaa Fadil, Yixuan He, Weijun Luo, Satoshi Koyama, Kristin Tsuo, Kaavya Paruchuri, Zhi Yu, Leland E. Hull, Zhili Zheng, Caitlin E. Carey, Raymond K. Walters, Benjamin M. Neale, Elise B. Robinson, Peter Kraft, Pradeep Natarajan, Alicia R. Martin

**Affiliations:** Analytic and Translational Genetics Unit, Massachusetts General Hospital, Boston, MA 02114, USA; Stanley Center for Psychiatric Research, Broad Institute of MIT and Harvard, Cambridge, MA 02142, USA; Program in Medical and Population Genetics, Broad Institute of MIT and Harvard, Cambridge, MA 02142, USA; Heart and Vascular Institute, Massachusetts General Brigham, Boston, 02114, MA, USA; Cardiovascular Disease Initiative, Broad Institute of MIT and Harvard, 415 Main St, Cambridge, MA 02142; Division of Medical Sciences, Harvard University, Boston, MA, 02115, USA; Human Genetics Center, The University of Texas Health Science Center at Houston School of Public Health, Houston, TX, USA; Department of Epidemiology, The University of Texas Health Science Center at Houston School of Public Health, Houston, TX, USA; College of Computing, The Georgia Institute of Technology, GA, 30332, USA; Laboratory for Cardiovascular Genomics and Informatics, RIKEN Center for Integrative Medical Sciences, Yokohama, Japan; Clinical and Translational Epidemiology Unit, Massachusetts General Hospital, Boston, MA 02114, USA; Division of General Internal Medicine, 100 Cambridge Street, Massachusetts General Hospital, Boston, MA, 02114, USA; Department of Medicine, Harvard Medical School, 25 Shattuck Street, Boston, MA 02115, USA; Division of Cancer Epidemiology and Genetics, National Cancer Institute, National Institutes of Health, Rockville, MD, USA; Center for Genomic Medicine, Massachusetts General Hospital, Boston, MA, 02114, USA

## Abstract

Polygenic scores (PGS) are typically derived from single-trait genome-wide association studies (GWAS), yet many complex diseases arise from shared genetic liability distributed across correlated clinical dimensions. Accordingly, disease risk depends not only on how genetic liability is represented but also on the social context in which that liability is expressed. Whether phenome-derived latent factors improve prediction, and how social determinants of health (SDoH) modify the realized utility of PGS, remains unclear.

Here we constructed PGS for 35 orthogonal latent phenomic factors derived from 2,772 phenotypes in 361,114 UK Biobank (UKB) participants and evaluated their phenomic specificity, cross-dataset portability and predictive performance relative to conventional disease-specific PGS across the UKB holdout, Mass General Brigham Biobank and the All of Us (AoU) Research Program. Factor-based PGS showed widespread, biologically coherent phenome-wide associations that were reproducible across biobanks and ancestries. Their predictive utility, however, was strongly disease dependent. For asthma, a respiratory factor PGS outperformed an internally derived disease-specific PGS and showed superior cross-ancestry portability, retaining 41.5% of European-ancestry predictive accuracy in African-ancestry individuals, compared with 22.9% for an asthma PGS derived from the largest available multi-ancestry GWAS. By contrast, disease-specific PGS remained superior for coronary artery disease (CAD) and type 2 diabetes (T2D). These findings suggest that phenome-derived aggregation is most beneficial when disease-specific GWAS incompletely capture underlying liability, including settings of biological heterogeneity or imprecise phenotyping.

We then evaluated SDoH in AoU as a complementary axis shaping prevalent disease prediction beyond genetic susceptibility. Across all three diseases, SDoH contributed substantial and largely independent predictive information beyond the disease-optimal genetic model. SDoH also modified how genetic liability translated into observed disease prevalence: for asthma and CAD, genetic stratification attenuated with increasing social burden, whereas this attenuation was substantially weaker for T2D. As a result, the same genetic percentile corresponded to different standardized predicted prevalences across social strata, reflecting disease-specific shifts in baseline prevalence, genetic gradients and calibration.

Together, these findings indicate that disease risk is shaped by both genetic liability and the social context in which that liability is realized. Phenome-derived PGS improve prediction under specific architectural conditions, whereas social context independently modifies the performance, calibration and interpretation of genetic risk across populations.

## Introduction

Polygenic scores (PGS) quantify inherited susceptibility to complex traits and diseases by aggregating genetic effects identified through genome-wide association studies (GWAS). Most PGS are derived from GWAS of a single observed phenotype, such as disease diagnoses or quantitative measurements. While effective for targeted outcomes, single-phenotype PGS may incompletely capture the underlying liability of complex diseases, especially when disease definitions are heterogeneous, noisy or sensitive to ascertainment. This limitation is particularly pronounced for complex diseases that lie along continuous spectra rather than discrete binary states. Data-driven representations that integrate information across correlated measurements may therefore provide more stable targets for genetic prediction^1,2^.

Modern population biobanks enable unprecedented phenotyping scales, collecting thousands of correlated clinical, behavioral, and socio-environmental measures for each participant. This phenotypic richness reveals structured covariance across traits, reflecting shared biological pathways, common disease processes and socially patterned exposures. Modeling each phenotype independently for PGS construction therefore overlooks this structure and treats correlated manifestations of disease as separate outcomes. Several approaches have sought to incorporate information from multiple traits for genetic prediction. Some combine phenotype-specific PGS downstream using linear or penalized regression models^3,4^. Others, including multi-trait GWAS methods such as MTAG^5^, jointly analyze genetically correlated traits to improve association estimates for selected outcomes. Although these approaches can improve prediction, they remain anchored to predefined observed phenotypes or prespecified target outcomes. They combine information across traits without redefining the unit of genetic signal itself.

Phenome-wide factor analysis provides an alternative framework by distilling thousands of heterogeneous phenotypes into a small number of latent dimensions that capture stable and genetically coherent axes of phenomic variation^6^. GWAS of factor scores thus shifts the unit of genetic association from individual phenotypes to latent phenomic dimensions. Because factor structures are cohort-specific and not directly transferable across datasets, however, it remains unknown whether genetic liabilities derived from such factors can generalize across populations and improve prediction of clinically relevant diseases relative to conventional disease-specific PGS.

A second limitation of polygenic prediction is that its performance depends not only on how genetic liability is represented, but also on the context in which that liability is expressed. PGS performance varies across contexts, such as age, socioeconomic status and other environmental settings^7–9^. Yet, these contextual differences are typically described post hoc rather than modeled explicitly. Social determinants of health (SDoH)^10^ provide structured and measurable representations of such context, capturing dimensions of social and environmental risk that influence disease burden and may modify the expression, interpretation and calibration of inherited susceptibility. Despite extensive epidemiological evidence linking social exposures to health outcomes, most polygenic prediction frameworks incorporate few explicit SDoH measures, limiting their ability to characterize how genetic risk manifests across heterogeneous environments^9,11,12^.

Here, we evaluate phenome-derived factor-based PGS as genetic representations of latent phenomic liability and assess their predictive utility across diseases and populations. Using factor scores derived from 361,114 deeply phenotyped UK Biobank (UKB) participants, we construct PGS for 35 orthogonal latent factors and benchmark their phenome-wide specificity, cross-cohort generalizability and predictive performance against disease-specific PGS. We then integrate structured SDoH indices in the All of Us Research Program (AoU), where rich survey-derived social measures enable explicit evaluation of social context beyond the disease-optimal genetic model, and test whether social context reshapes the performance, calibration and interpretation of genetic risk. Through this integrated framework, we define when phenome-derived genetic representations improve disease prediction, and show that social context independently modifies the realized meaning of genetic risk.

## Results

Previous phenome-wide factor analysis in 361,114 unrelated European (EUR) individuals in UKB identified 35 latent factors distilled from 2,772 heterogeneous phenotypes^6^. These factors spanned 6 broad domains: clinical disease and health history, lifestyle and behavior, mental health and cognition, physical measurements, socio-environmental context and biomarkers (**Table S1**). Using GWAS of factor scores, we constructed PGS for each factor with PRS-CS^13^, yielding genetic predictors of latent phenomic liability. Because latent factor scores are cohort-specific and cannot be reconstructed outside the discovery cohort, we assessed factor-based PGS through phenome-wide association analyses and predictive benchmarking across multiple biobanks and ancestry groups. As a complementary component of the study, we evaluated SDoH in AoU using structured indices derived from 29 variables spanning the five Healthy People 2030 domains^10^. We constructed both domain-specific scores and a composite SDoH index, then examined their independent and joint contributions beyond the disease-optimal PGS, as well as whether SDoH burden modified the performance, calibration, and interpretation of genetic risk through interaction and stratified analyses. The overall analytical framework is summarized in **Fig. 1**.

**Figure 1.**
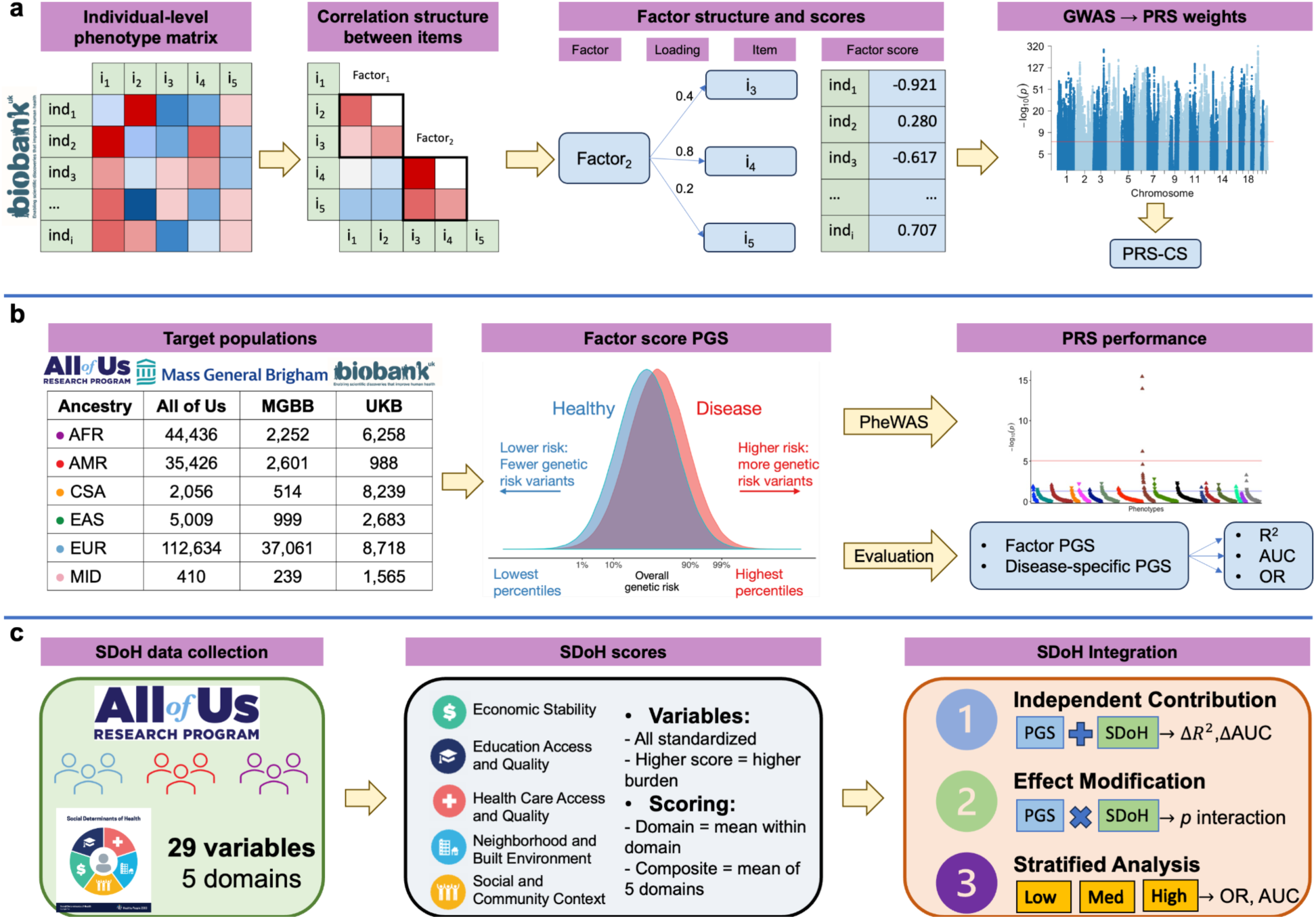
Study design for evaluating phenome-derived factor-based polygenic scores across biobanks. **a)** Previous phenome-wide factor analysis in 361,114 unrelated individuals of European ancestry in the UK Biobank (UKB) identified 35 latent factors distilled from 2,772 heterogeneous phenotypes, as described by Carey et al^6^. Using GWAS summary statistics for each factor, we constructed factor-based polygenic scores (PGS) with PRS-CS. **b)** Factor-based PGS were evaluated in independent individuals from three large biobanks: a UKB holdout set, the Mass General Brigham Biobank (MGBB), and the All of Us Research Program (AoU), spanning multiple ancestry groups. We assessed phenomic specificity and generalizability using PGS-based phenome-wide association studies (PGS-PheWAS) and evaluated predictive performance using variance explained (*R*^2^), area under the receiver operating characteristic curve (AUC), and odds ratio (OR), with comparisons to conventional disease-specific PGS derived from case-control GWAS. AFR: African; AMR: Admixed American; CSA: Central and South Asian; EAS: East Asian; EUR: European and MID: Middle Eastern. **c)** Social determinants of health (SDoH) were evaluated in AoU using 29 variables spanning the five Healthy People 2030 domains. Variables were standardized and aggregated into domain-specific scores and a composite SDoH index, with higher values indicating greater social burden. We then assessed three aspects of genetic-social interplay: the independent contribution of SDoH beyond the disease-optimal PGS, effect modification through multiplicative PGS×SDoH interaction models, and context-dependent calibration and stratification across SDoH burden tertiles. See **Methods** for additional details.

### Factor-based PGS capture specific and reproducible phenomic structure across biobanks and ancestries

To assess the phenomic specificity and generalizability of factor-based PGS, we conducted PGS-pheWAS association analyses within each biobank-by-ancestry stratum, applying Bonferroni correction based on the number of tested outcomes per stratum (**Methods**). Across biobanks, we observed widespread and reproducible phenomic associations of factor-based PGS. Specifically, we identified a total of 96, 698, and 3,869 significant PGS-PheCode associations out of 42,245, 115,253, and 190,785 tests in the UKB holdout, Mass General Brigham Biobank (MGBB), and AoU, respectively (**Table S2)**. Because factor models are invariant to sign, factor-PGS were interpreted relative to dominant loadings defining each factor, such that positive and negative associations denote concordant and discordant relationships with the corresponding latent phenomic dimension. Under this convention, the majority of significant associations were positive (84.9%), with a median OR of 1.12, while negative associations showed a median OR of 0.91. The predominance of modest, directionally coherent effects across biobanks is consistent with highly polygenic liability rather than cohort-specific artifacts. Both positive and negative associations were biologically interpretable in the context of the constituent phenotypes defining each factor.

The distribution of associations across factors revealed clear and structured phenomic structure (**Extended Data Figs. 1-2**). Factors reflecting clinical disease and health history showed the highest number of associations (2,199), followed by socio-environmental contexts (964) and physical measurements (858). These patterns were consistent across biobanks despite substantial differences in recruitment strategies, healthcare systems, and phenotype ascertainment, indicating that factor-based PGS capture robust dimensions of genetic liability.

Factor-based PGS further exhibited strong phenotypic specificity consistent with the domains captured by their constituent phenotypes. For example, the PGS derived from a respiratory disease factor showed strong and selective associations with PheCodes within the respiratory disease category across biobanks and ancestries. Asthma emerged as the top association in each dataset, with ORs ranging from 1.32 in AoU and MGBB to 1.57 in the UKB holdout among EUR individuals (**Fig. 2**). Consistent with domain specificity, the respiratory factor PGS showed minimal enrichment for associations outside respiratory-related PheCode chapters (**Table S2**), indicating that phenome-wide signal was concentrated within clinically coherent domains rather than reflecting diffuse pleiotropy. In contrast, PGS derived from socio-demographic factors, such as educational attainment, exhibited broader phenomic associations spanning multiple disease categories, with ORs ranging from 0.84 to 1.70 (**Fig. 2)**. These associations were not confined to a single disease domain and included consistent enrichment for mental and behavioral disorders across biobanks. This broader pattern likely reflects a combination of shared behavioral risk profiles and socially patterned exposures, rather than a single underlying biological process.

**Figure 2.**
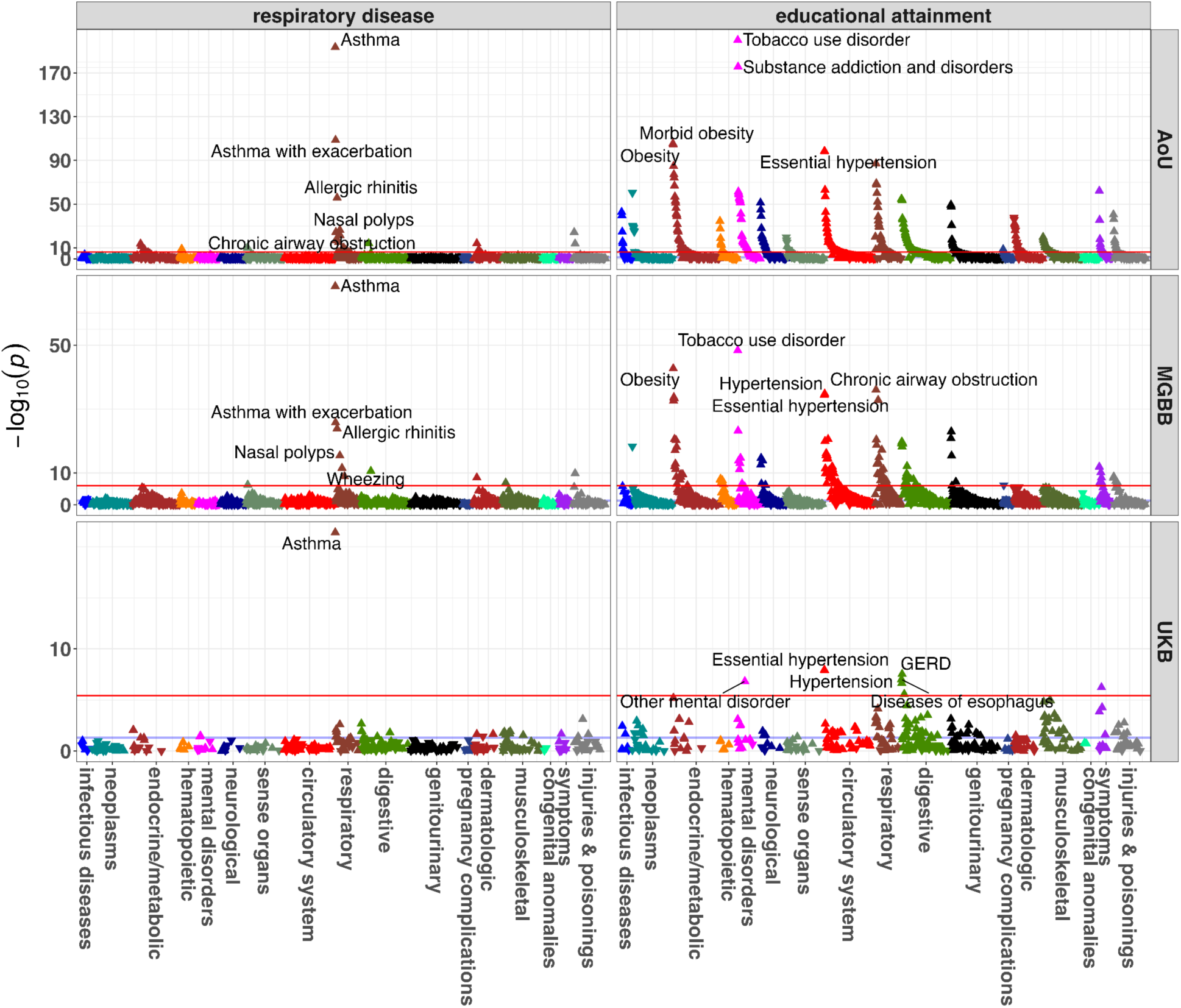
PGS-pheWAS associations in European populations across biobanks. The blue line indicates nominal significance (*p* < 0.05) and the red line indicates the Bonferroni corrected significance threshold. We illustrated the results in European populations using PGS derived from respiratory disease and educational attainment factors as examples. We annotated the top 5 significant associations in each biobank. The full results are detailed in **Table S2**.

To quantify phenomic structure at the level of disease categories, we performed enrichment analyses assessing whether significant associations for each factor were disproportionately concentrated within specific PheCode chapters. Within each biobank and ancestry, enrichment *Z*-scores were computed using a hypergeometric framework that accounted for the number of tested outcomes per factor and category (**Methods; Supplementary Note 1**). Meta-analysis of enrichment statistics across biobanks revealed highly coherent and biologically interpretable patterns (**Fig. 3; Extended Data Fig. 3**). Disease-oriented factors showed strong positive enrichment for their corresponding clinical domains, particularly in EUR (median meta-analysis Z = 15.22; range 3.80-21.22), with consistent but attenuated signals observed across other ancestries. In contrast, socio-demographic and behavioral factors demonstrated broader but generally weaker enrichment across multiple disease categories (median Z = 3.74; range 2.43-17.86 in EUR), consistent with more diffuse cross-domain effects.

**Figure 3.**
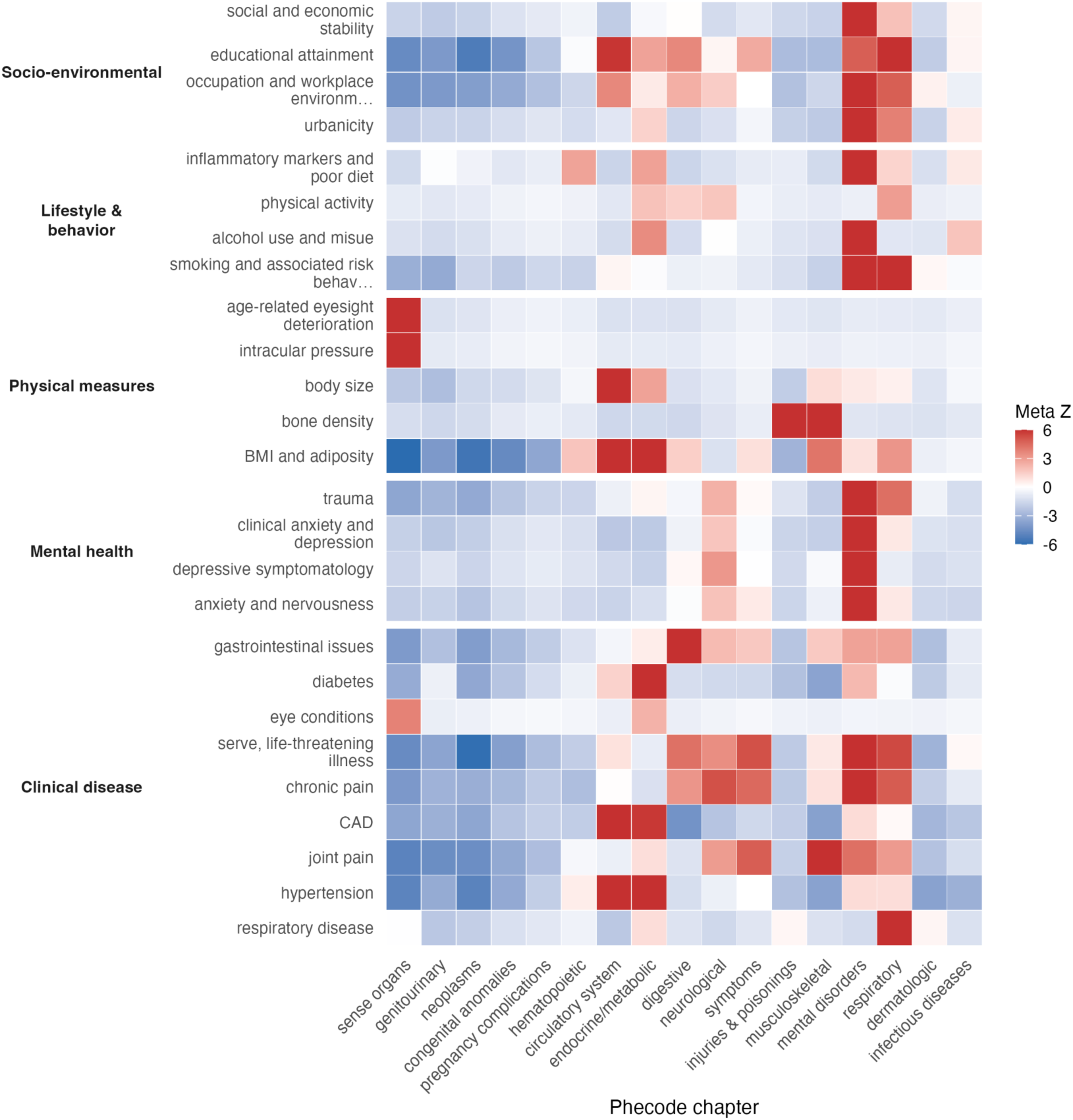
Meta-analysis of phenomic enrichment in European ancestry reveals structured and interpretable genetic liability across disease domains. Heatmap shows meta-analytic enrichment Z-scores summarizing the concentration of significant factor-based PGS associations within PheCode chapters across biobanks in individuals of European ancestry (EUR). Columns correspond to latent phenomic factors grouped into six broad domains, and rows correspond to PheCode chapters. Enrichment statistics were meta-analyzed across biobanks using Stouffer’s method, and only significant associations identified in at least two biobanks are shown (see **Methods**). Color indicates the direction and magnitude of enrichment (Meta Z), with red denoting positive enrichment and blue denoting negative enrichment. White indicates little or no enrichment (Z ≈ 0). PheCode chapters (x-axis) were ordered by hierarchical clustering of enrichment Z-scores across factors.

Importantly, these enrichment patterns were consistent across biobanks despite the non-transferability of the underlying phenotypic factor structures (**Table S3; Supplementary Figs. 1-3**). Although latent factors derived from deep phenotyping cannot be directly reconstructed in external datasets, the associated liabilities manifested as stable and expected phenomic signatures across independent populations–an importantly portable characteristic of phenome-derived PGS.

Across ancestries, we observed strong concordance in the direction and relative magnitude of associations. Among all significant associations, median ORs were 1.11 (s.d. = 0.13) across 411 associations in EUR, 1.19 (s.d. = 0.17) across 298 associations in Admixed American ancestry (AMR), 1.15 (s.d. = 0.08) across 61 associations in individuals of African ancestry (AFR), 1.30 (s.d. = 0.13) across 38 associations in Central and South Asian ancestry (CSA), and 1.52 (s.d. = 0.17) across 9 associations in East Asian ancestry (EAS). An additional two associations were observed in individuals of Middle Eastern ancestry. Among these, 411 associations were shared by at least two ancestry groups, and effect sizes were strongly correlated across ancestry pairs with adequate overlap (Pearson’s *r* = 0.80-0.89). Effect-size distributions were similarly comparable across biobanks, with median ORs of 1.31 (s.d. = 0.21) in the UKB holdout, 1.11 (s.d. = 0.13) in All of Us, and 1.12 (s.d. = 0.13) in MGBB, and 640 associations were replicated in at least two biobanks. Overall, 50 associations involving 10 distinct factors and 28 PheCodes were detected across all three biobanks in at least one ancestry group (**Extended Data Fig. 4**).

Together, these analyses indicate that factor-based PGS capture stable and interpretable dimensions of genetic liability that manifest as reproducible phenomic signatures across independent biobanks and populations.

### Factor-based PGS show disease-dependent predictive performance

Having established phenomic specificity and cross-dataset generalizability of factor-based PGS, we next evaluated their utility for disease prediction. We first benchmarked factor-based PGS against internally derived PheCode-based disease PGS from matched GWAS in the same discovery samples used for factor GWAS, enabling a direct comparison under matched ascertainment. As a secondary benchmark, we compared these results with disease-specific PGS derived from large-scale public case-control GWAS (**Table S4**). Candidate outcomes were identified empirically on the basis of replicated PGS-pheWAS associations across biobanks and concordance between factor composition and top disease associations (**Methods**). From this set, we selected asthma, coronary artery disease (CAD) and type 2 diabetes (T2D) as primary outcomes for detailed benchmarking. We quantified their predictive performance using incremental variance explained on the liability scale (*R*^2^) relative to the covariate-only model (**Methods)**. Under matched internal benchmarking, factor-based PGS performed better for asthma, whereas disease-specific PGS consistently dominated for cardiometabolic conditions (**Fig. 4; Table S5**). By contrast, external disease-specific PGS derived from large public GWAS generally outperformed factor-based PGS across all three diseases (**Extended Data Figs. 5 and 6**).

**Figure 4.**
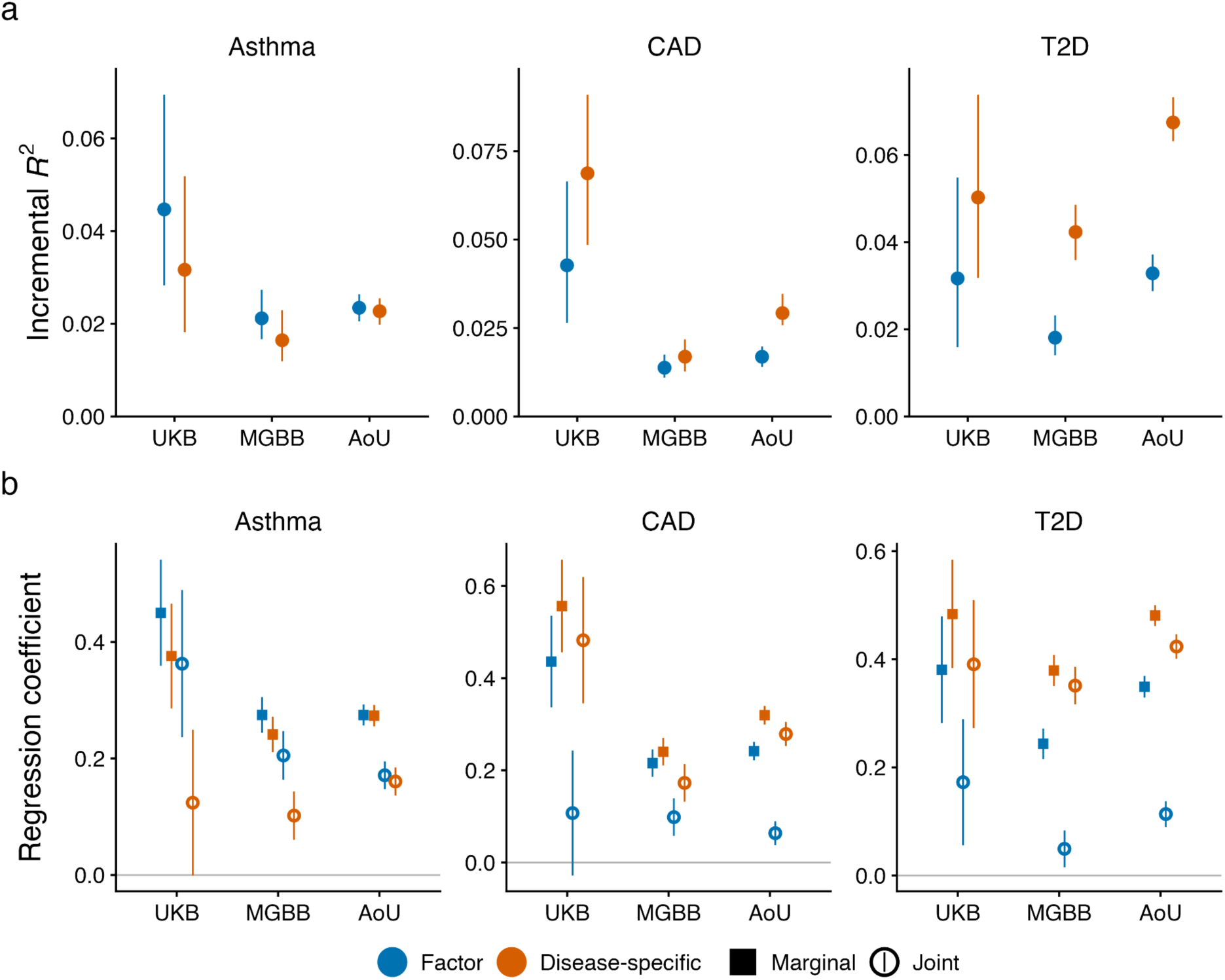
Disease-dependent predictive performance of factor-based and disease-specific polygenic scores across biobanks. **a)** Predictive accuracy of factor-based and disease-specific (PheCode-based) PGS for asthma, coronary artery disease (CAD), and type 2 diabetes (T2D) across the UK Biobank (UKB) holdout set, Mass General Brigham Biobank (MGBB), and the All of Us Research Program (AoU) in European-ancestry individuals. Points indicate incremental *R*^2^ on the liability-scale, and error bars indicate 95% confidence intervals. **b)** Standardized regression coefficients for factor-based and disease-specific PGS estimated in marginal models (each PGS evaluated separately) and joint models (both PGS included simultaneously). Points indicate coefficient estimates and error bars indicate 95% confidence intervals.

#### Factor-based PGS outperform disease-specific scores for asthma

Asthma showed the strongest improvement in predictive performance for factor-based PGS. Across EUR cohorts, the respiratory factor PGS performed comparably to or better than the internally derived PheCode-based asthma PGS. The largest improvement was observed in UKB, where factor PGS achieved *R*^2^ = 0.045 (95% CI: 0.028-0.069) compared to 0.032 (0.018-0.052) for asthma PGS, corresponding to a 41% relative gain (**Fig. 4a**; **Table S5**). This advantage replicated in MGBB (+29%) and was near parity in the largest cohort, AoU (+3%).

The two PGS were moderately to strongly correlated across cohorts (Pearson’s *r* = 0.67-0.70), indicating substantial shared genetic liability. However, joint models including both scores revealed that factor PGS captured independent predictive signals beyond asthma PGS (**Fig. 4b; Table S5)**. In UKB, factor PGS remained strongly associated with asthma (β = 0.363, p = 1.9×10⁻⁸), while asthma PGS became marginally non-significant (β = 0.124, p = 0.052). In MGBB, both scores contributed independently, though the effect size of factor PGS was approximately two-fold larger. In AoU, both scores contributed comparably, consistent with balanced predictive signal in the largest cohort.

As a secondary benchmark, we compared the respiratory factor PGS with an asthma PGS derived from the Global Biobank Meta-analysis Initiative (GBMI)^14,15^. Against this external benchmark, factor PGS showed comparable performance in UKB (*R*^2^ = 0.045 vs 0.050), whereas GBMI PGS exhibited increasing advantage in MGBB (50% higher) and AoU (77% higher), consistent with its substantially larger discovery sample size (**Extended Data Fig. 5**). Despite this, factor PGS showed markedly superior cross-ancestry portability, even though its underlying GWAS were conducted solely in EUR participants, whereas the GBMI meta-analysis included multiple ancestries. In AFR, factor PGS retained 41.5% of EUR predictive accuracy, compared to 22.9% for GBMI PGS and 12.1% for the smaller internal PheCode-based PGS (**Extended Data Fig. 6**). Similar advantages were observed in EAS and CSA, with comparable performance in AMR.

This predictive advantage was accompanied by differences in underlying genetic architecture. The factor GWAS exhibited significantly higher SNP heritability than PheCode-based asthma GWAS (ℎ^2^ = 0.099 ± 0.009 vs 0.044 ± 0.004; *p* < 0.001), while the two GWAS remained highly genetically correlated (*r*_g_ = 0.81 ± 0.02). MAGMA pathway analysis further revealed a substantially richer and stronger pathway enrichment for factor GWAS, with 224 significantly enriched pathways (FDR < 0.05), dominated by type-2 immune and eosinophil-mediated processes, compared to 123 pathways for asthma GWAS with weaker enrichment signals (**Methods**; **Supplementary Fig. 4**). Together, these findings suggest that the respiratory factor PGS captures a broader, more biologically coherent, and a more portable genetic representation of asthma-related liability.

#### Disease-specific PGS outperform factor-based scores for cardiometabolic diseases

In contrast to asthma, disease-specific PGS consistently outperformed factor-based PGS for CAD and T2D across cohorts. For CAD, disease-specific PGS showed higher predictive accuracy in all three cohorts (**Fig. 4**). In UKB, CAD PGS achieved *R*^2^ = 0.069 (0.048-0.091) compared with 0.043 (0.026-0.066) for the corresponding factor PGS (+60%). This advantage remained larger in AoU (+73%) but was attenuated in the clinical cohort MGBB (+21%). The two scores were highly correlated (*r* ≈ 0.68), reflecting substantial shared genetic architecture, yet joint modeling indicated little additional contribution from factor-based PGS. Although factor and CAD GWAS were highly genetically correlated (*r*_g_ = 0.88), pathway analysis revealed broader vascular and structural pathway involvement for CAD GWAS, while the corresponding factor GWAS was more narrowly enriched for lipoprotein and triglyceride-related processes (**Supplementary Fig. 4**). This pattern is consistent with the factor’s dominant loadings on ischemic heart disease diagnoses together with cardiovascular medications and high-cholesterol-related items. The same pattern was observed in secondary comparisons with large public CAD GWAS-based PGS (**Extended Data Figs. 5 and 6**).

A similar pattern was observed for T2D, with larger relative differences in predictive performance between disease-specific and factor-based PGS. In UKB, PheCode-based T2D PGS achieved *R*^2^ = 0.050 (0.032-0.074) compared to 0.032 (0.016-0.055) for factor PGS, representing a 56% relative improvement; larger relative differences were observed in MGBB (+130%) and AoU (+101%) (**Fig. 4; Table S4**). Although the two scores showed moderate correlation (*r* = 0.55-0.56), joint modeling indicated that T2D PGS captured most predictive signals, with factor PGS contributing minimal incremental value. These performance differences were concordant with underlying genetic architecture. T2D GWAS exhibited modestly higher heritability (ℎ^2^ = 0.052 ± 0.003 vs 0.043 ± 0.003; *p* = 0.03) and broader MAGMA enrichment (**Supplementary Fig. 4**), whereas the corresponding factor was dominated by diabetes diagnosis together with cholesterol-, medication-, and illness-burden-related items. Secondary benchmarking against large public T2D GWAS likewise favored disease-specific PGS (**Extended Data Figs. 5-6**).

Beyond the three primary outcomes, additional factor-disease pairs showing similar patterns in internal benchmarking were observed across other phenotypic domains. For example, the bone-density factor improved prediction of osteoporosis and related fracture outcomes, and the joint-pain factor showed improved prediction of degenerative spinal disease across multiple biobanks. However, these effects were generally smaller and less consistently replicated than those observed for asthma. Full results across all candidate outcomes are presented in **Extended Data Fig. 7**.

### Social determinants of health independently and contextually shape genetic prediction in AoU

Having identified the disease-optimal genetic representation for each outcome phenotype (factor-based PGS for asthma; disease-specific PGS for CAD and T2D), we next evaluated SDoH exclusively in AoU, where structured social measures were available at sufficient depth for joint modeling. We first focused on AoU-EUR, using a composite SDoH score derived from 29 variables spanning five Healthy People 2030 domains. These variables were collected across three AoU surveys, and missingness varied substantially by survey, with variable-level missingness ranging from 0% to 59% among EUR participants (**Methods; Table S6**). Unless otherwise noted, analyses in this section were performed under a prevalent-disease framework, including association, calibration, and stratified evaluation of genetic risk. We then performed incident-disease analyses as a prospective sensitivity analysis to evaluate whether the main predictive patterns extended to future disease risk.

#### Social determinants of health provide substantial and largely independent predictive information beyond genetic risk

In AoU-EUR, SDoH alone explained a substantial proportion of disease liability under this prevalent-disease framework (asthma: *R*^2^ = 0.023; CAD: *R*^2^ = 0.025; T2D: *R*^2^ = 0.048), with the largest contribution observed for T2D. These effects were comparable in magnitude to single genetic predictors (**Fig. 5**), indicating that social context captures a meaningful component of disease risk not encoded by inherited variation. Joint modeling further increased predictive performance (**Fig. 5a)**. For asthma, adding SDoH to the factor-based PGS increased variance explained to 0.046, approximately doubling predictive performance relative to the genetic model alone. For cardiometabolic disease, combining SDoH with disease-specific PGS yielded *R*^2^ = 0.050 for CAD and *R*^2^ = 0.104 for T2D. Both PGS and SDoH remained significantly associated in joint models (all *p* < 10^-100^), with concordant effects on the odds-ratio scale (**Table S7)**. Correlation between the optimal PGS and the SDoH composite was weak across diseases (Pearson’s *r* < 0.1), suggesting limited overlap between genetic and social predictors. Together with the joint-model results, this pattern indicates that social context captures a substantial and largely distinct component of disease liability beyond inherited variation.

**Figure 5.**
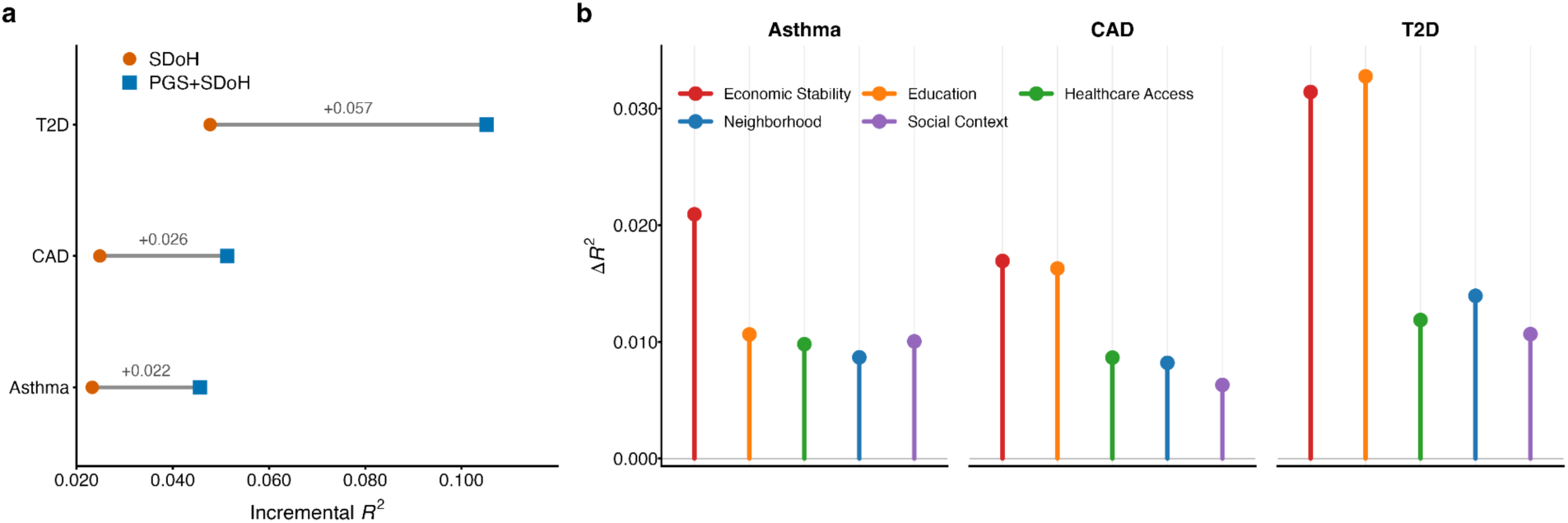
Social determinants of health provide substantial and largely independent predictive information beyond genetic risk. **a)** Incremental liability-scale variance explained (*R*^2^) by social determinants of health (SDoH) alone and by the joint PGS+SDoH model for asthma, CAD (coronary artery diseases) and T2D (type 2 diabetes). Grey line segments connect the two estimates for each disease and indicate the gain in predictive performance obtained by adding PGS to the SDoH-only model. **b)** Domain-specific incremental variance explained (*ΔR*^2^) when each SDoH domain was added individually to the optimal PGS model.

We next evaluated domain-specific contributions by adding each SDoH domain individually to the disease-optimal genetic model (**Fig. 5b**). For asthma, economic stability provided the largest incremental contribution (*ΔR*^2^ = 0.021). For CAD and T2D, economic stability and education contributed similarly and were the dominant SDoH domains (CAD: *ΔR*^2^ = 0.017 vs 0.016; T2D: *ΔR*^2^ = 0.033 vs 0.031). Healthcare access, neighborhood characteristics and social context contributed modest but consistent variance across diseases (*ΔR*^2^ = 0.006-0.013). The summed variance of individual domains exceeded that of the composite contribution, reflecting overlapping contributions rather than independent effects. This pattern supports the use of a composite SDoH index to summarize shared socio-environmental burden.

To reduce concern that SDoH associations might partly reflect downstream consequences of disease rather than antecedent exposures, we repeated analyses using a 16-variable temporally robust SDoH subset selected from questionnaire items judged to reflect relatively stable structural or socioeconomic conditions (**Methods; Table S6**). In AoU-EUR, the incremental contributions of SDoH beyond the disease-optimal genetic model were largely preserved across diseases using this reduced set, with liability-scale *ΔR*^2^ retention of 78% for asthma and approximately complete retention for CAD and T2D relative to the full 29-variable composite; all joint associations remained highly significant (**Table S7 and S8**). Because the primary analyses are based on prevalent disease, this sensitivity analysis does not establish temporal ordering, but it indicates that the main findings are not driven solely by short-term or illness-proximal SDoH measures.

As a prospective sensitivity analysis, we next evaluated incident disease in AoU-EUR using Cox models (**Methods**). SDoH remained strongly associated with incident disease in joint models, with hazard ratios per standard deviation of 1.39 for asthma, 1.42 for CAD, and 1.48 for T2D (**Extended Data Fig. 8a**). Adding SDoH to the genetic model also improved the 5-year AUC from 0.597 to 0.664 for asthma, from 0.768 to 0.794 for CAD, and from 0.684 to 0.742 for T2D (**Extended Data Fig. 8b**).

Together, these results show that SDoH contributes substantial and largely independent predictive information beyond the disease-optimal genetic model, both for prevalent disease burden and for future disease risk.

#### Social determinants of health systematically reshape the calibration and predictive value of genetic risk

Having established that SDoH contributes independent predictive information in AoU-EUR, we next examined whether the predictive meaning of a given level of genetic risk is consistent across social context. Specifically, we tested whether the mapping from genetic percentile to standardized predicted prevalence remains stable across SDoH strata, or whether SDoH burden shifts baseline prevalence, alters genetic stratification, and changes calibration in a disease-specific manner. To address this, we evaluated interaction models, stratified predictive performance, standardized predicted prevalence across joint strata of genetic and social burden, and calibration across SDoH burden tertiles (**Methods**).

In AoU-EUR, stratified analyses showed disease-dependent attenuation of PGS predictive performance across SDoH burden tertiles (**Fig. 6a**; **Extended Data Fig. 9; Table S9**). For asthma, the incremental AUC (*Δ*AUC) contributed by the disease-optimal PGS declined markedly with increasing SDoH burden, from 0.038 in the lowest tertile to 0.018 in the highest. CAD showed a smaller decline, from 0.011 to 0.006, whereas T2D showed minimal change, decreasing slightly from 0.035 to 0.031. Risk stratification on the odds-ratio (OR) scale showed broadly concordant attenuation for asthma and CAD (**Table S10; Extended Data Fig. 10a**). Comparing individuals in the top versus bottom PGS decile, ORs decreased from 2.96 (95% CI: 2.48-3.54) to 2.43 (2.10-2.81) for asthma and from 3.21 (2.70-3.82) to 2.52 (2.10-3.02) for CAD across increasing SDOH burden tertiles. By contrast, T2D did not show the same monotonic attenuation pattern across SDoH tertiles, with corresponding ORs of 4.43, 5.10 and 4.81. Multiplicative interaction tests nevertheless identified significant negative PGS×SDoH interactions for all three diseases (**Table S7**), indicating attenuation of genetic effects on the continuous scale even when tertile-based contrasts were weak or non-monotonic. Similar patterns were observed when stratifying per-standard-deviation PGS effects across SDoH tertiles (**Extended Data Fig. 10b**).

**Figure 6.**
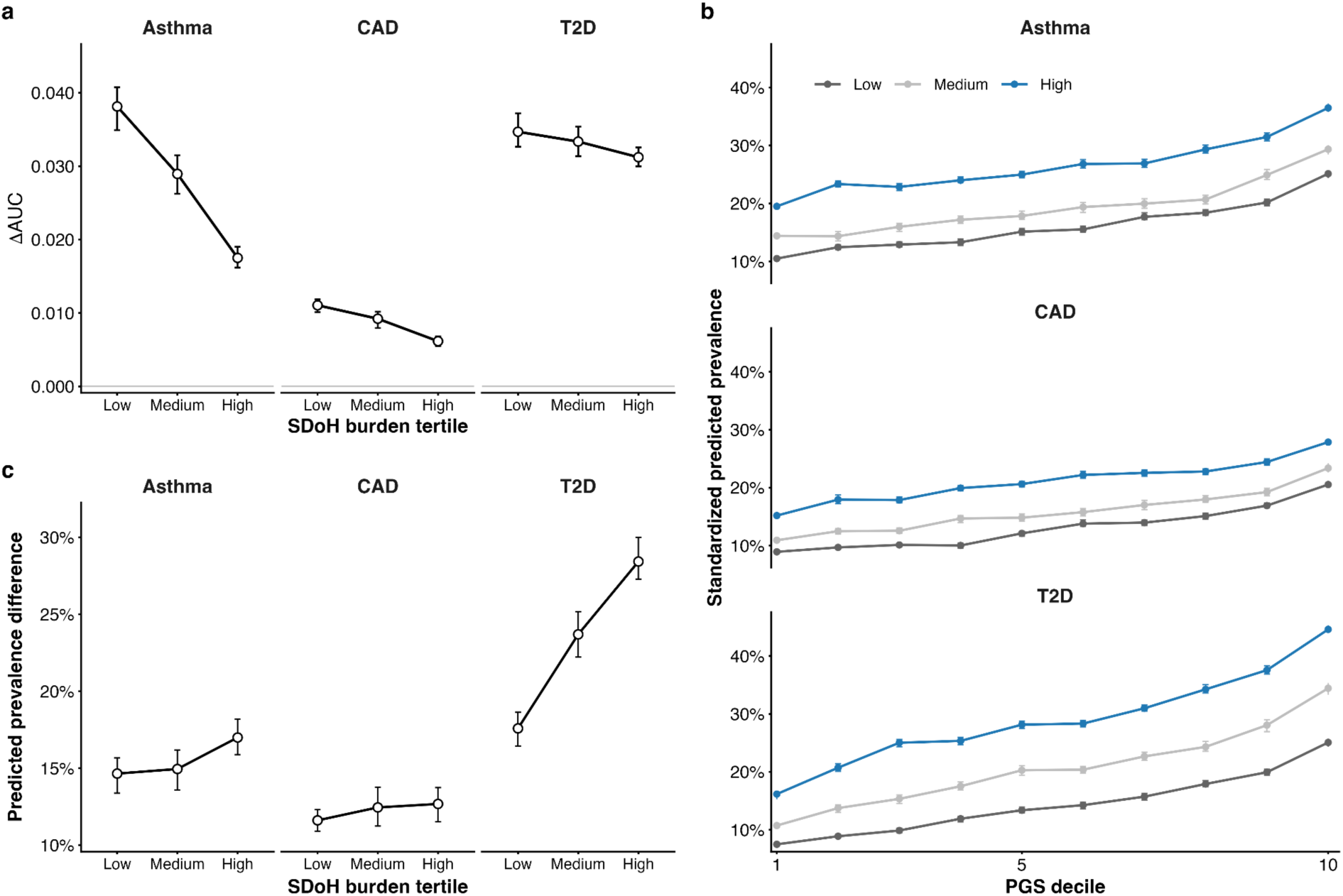
Social context reshapes the realized meaning of genetic risk in a disease-specific manner. **a**) Incremental predictive performance contributed by the optimal polygenic score (PGS), measured as *Δ*AUC relative to covariates-only model, stratified by tertiles of social determinants of health (SDoH) burden (Low, Medium, High) for asthma, coronary artery disease (CAD) and type 2 diabetes (T2D) in AoU-EUR. Points indicate estimates and error bars indicate 95% confidence intervals. **b**) Standardized predicted prevalence across PGS deciles within each SDoH tertile, obtained by averaging fitted probabilities over the empirical covariate distribution of the analysis sample (**Methods**). Lines connect decile-specific estimates for each SDoH stratum, illustrating joint effects of genetic and social risk on disease prevalence. **c**) Predicted prevalence difference between the highest and lowest PGS deciles (decile 10 minus decile 1) within each SDoH tertile. Points denote estimates and error bars indicate 95% confidence intervals.

To interpret how these interactions manifest on the standardized predicted-prevalence scale, we estimated age-standardized predicted prevalence across joint strata of genetic and social burden (PGS decile × SDoH burden tertile; **Fig. 6b**), using model-based predictions averaged over the empirical covariate distribution of the analysis sample (**Methods**). In AoU-EUR, SDoH shifted baseline predicted prevalence even among individuals in the lowest PGS decile. For asthma, standardized predicted prevalence increased from 0.105 to 0.195 across SDoH tertiles; for CAD, it increased from 0.089 to 0.152; and for T2D, from 0.075 to 0.162. Thus, SDoH burden shifted baseline disease prevalence independently of genetic predisposition under this standardized prediction framework.

The relationship between genetic percentile and standardized predicted prevalence also differed systematically across social strata (**Fig. 6c**). For asthma, the predicted prevalence difference between the highest and lowest PGS deciles increased modestly with SDoH burden, from 0.147 to 0.170. T2D showed a much larger increase, from 0.176 to 0.284, indicating stronger separation of genetic risk groups under greater social burden. By contrast, CAD showed only limited change after age standardization, from 0.116 to 0.127, indicating weaker context dependence on the predicted-prevalence scale than suggested by the predictive performance analyses. These results show that identical genetic percentiles do not correspond to a fixed predicted prevalence across social environments.

In AoU-EUR, calibration analyses likewise showed that the mapping from genetic percentile to predicted prevalence varied systematically across social strata (**Extended Data Fig. 11; Table S11**). Calibration intercepts shifted monotonically with SDoH burden for all three diseases, reflecting relative overprediction in lower-SDoH strata and underprediction in higher-SDoH strata. For example, asthma calibration intercepts shifted from −0.399 to −0.187 to +0.456 across SDoH tertiles, with similar directional patterns observed for CAD and T2D. Calibration slopes varied more modestly, indicating that SDoH primarily shifted baseline prevalence and calibration-in-the-large, while only partially altering the scaling of genetic risk. Tail analyses reinforced this non-comparability across social context. Among individuals in the highest PGS decile, standardized predicted prevalence increased across SDoH tertiles for asthma and T2D (**Fig. 6b**), indicating that the same level of genetic risk corresponded to different realized disease prevalence across social strata. CAD showed a smaller upward shift, consistent with its weaker context dependence on the standardized predicted-prevalence scale.

Age- and sex-stratified analyses showed directionally concordant interaction effects across strata, although statistical support varied by diseases: signals were most evident in females and individuals aged 50-65 years for asthma, in males and those aged 50 years or older for CAD, and in both sexes but primarily below age 65 for T2D (**Table S12**).

Together, these findings indicate that models omitting SDoH are not uniformly calibrated across social environments, but instead exhibit structured miscalibration that changes the predictive meaning of genetic risk in a disease-specific manner.

#### Cross-ancestry generalizability under a harmonized SDoH subset

To evaluate whether the independent and largely orthogonal contributions of genetic and social risk generalize beyond AoU-EUR, we constructed a harmonized SDoH composite comprising seven socioeconomic indicators from the AoU Basics Survey with low missingness (<1%) across populations (**Methods; Table S6**). This smaller set of variables did not include measures of social and community context (n=4), some measures of neighborhood and built environment (n=4), some measures of healthcare quality and access (n=9), and some measures of economic stability (n=5). In EUR, the reduced composite retained 58%, 80% and 95% of the variance explained by the full 29-variable score for asthma, CAD and T2D, respectively, indicating attenuation due to reduced domain coverage but preservation of most predictive signal for cardiometabolic outcomes. Comparison of the full and harmonized composites within EUR showed that attenuation under the reduced score was concentrated in the SDoH component, whereas optimal-PGS coefficients in joint models were largely unchanged (**Table S13**).

We then applied the harmonized composite within AoU in AFR and AMR populations to evaluate cross-ancestry robustness under constrained SDoH measurement. Adding SDoH to ancestry-specific optimal PGS increased liability-scale variance explained, with smaller gains in AFR (0.001-0.004) than in AMR (0.004-0.014) and EUR (0.014-0.037) (**Fig. 7; Table S14**). Joint-model effect estimates showed that attenuation was primarily driven by the SDoH component rather than the genetic component. In EUR and AMR, both the harmonized SDoH composite and the optimal PGS remained positively associated across all three diseases. In AFR, the PGS term remained significant for all diseases, whereas the SDoH term was attenuated and no longer significant for CAD and T2D. Relative to EUR, SDoH coefficients were reduced to approximately 58-76% in AMR and 15-53% in AFR, whereas PGS coefficients were largely preserved (**Supplementary Fig. 5**). These patterns indicate preservation of the main SDoH signal in AMR, with substantially greater attenuation in AFR. Within AoU, correlation between the harmonized SDoH composite and ancestry-specific PGS remained low across populations (Pearson’s *r* = 0.007-0.012 for asthma, 0.020-0.068 for CAD and 0.036-0.091 for T2D), again supporting largely distinct contributions from genetic and social predictors.

**Figure 7.**
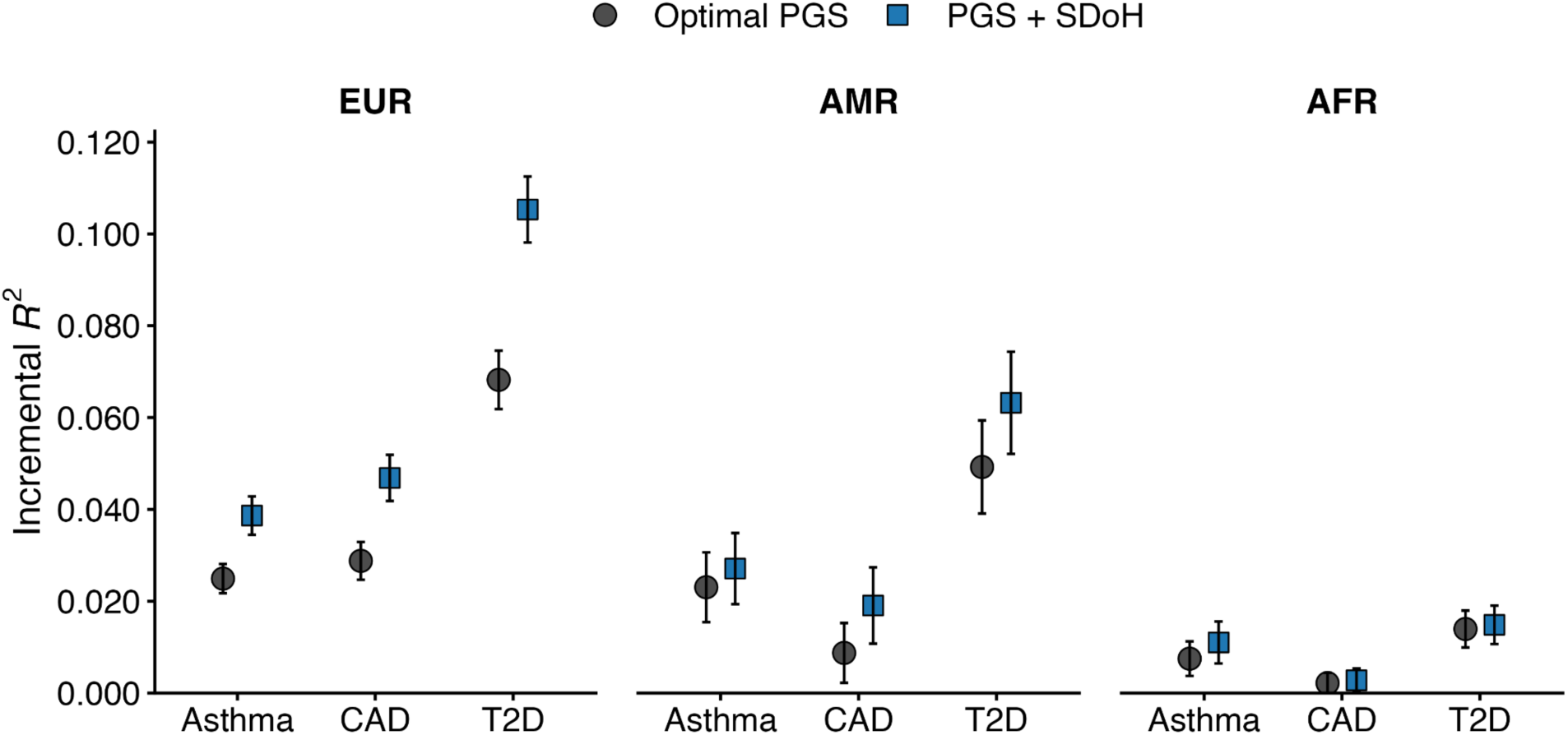
Cross-ancestry generalizability of joint genetic-social prediction under a harmonized SDoH composite. Incremental variance explained (*R*^2^) on the liability scale for the optimal PGS and the joint PGS+SDoH model for asthma, coronary artery disease (CAD) and type 2 diabetes (T2D), stratified by African (AFR), Admixed American (AMR) and European (EUR) ancestry groups. Points denote point estimates and error bars indicate 95% confidence intervals. The joint model incorporated the harmonized 7-variable social determinants of health (SDoH) composite.

Predicted prevalence and calibration analyses under the reduced composite showed socially patterned shifts in baseline prevalence and calibration across SDoH strata in EUR and AMR populations, broadly consistent with the EUR patterns under the full SDoH representation (**Supplementary Fig. 6; Table S14 and S15**). In AFR, these patterns were weaker and less consistent, in line with attenuation of the harmonized SDoH component in joint models. These results indicate that the predictive meaning of a given genetic percentile varies across social environments beyond EUR, although the magnitude and consistency of this effect are attenuated under the reduced SDoH representation.

Exploratory interaction analyses under the harmonized composite showed consistently negative interaction estimates across ancestries, with the strongest statistical support in EUR and for T2D in AMR (**Fig. S7**). To assess whether integrating SDoH improved cross-population generalizability of prediction, we compared the relative retention of predictive performance across ancestries for PGS-only and joint PGS+SDoH models (**Table S16**). Adding SDoH did not consistently improve non-EUR retention relative to EUR: attenuation was observed across all three diseases in AFR, and modest improvement was seen only for CAD in AMR. These results indicate that, under the harmonized 7-variable representation, cross-ancestry limitations of the joint model reflect reduced transferability of the SDoH component more than loss of genetic signal.

Collectively, these analyses show that the independent contribution of social context and its modulation of genetic risk extend beyond EUR, but also highlight that cross-ancestry calibration remains constrained by incomplete harmonization of SDoH measurement. More broadly, these findings should be interpreted as estimates within the AoU setting, where both the distribution and measurement of social context may differ from those in other healthcare systems and populations.

## Discussion

This study shows that the utility of polygenic prediction depends on two related but distinct features of disease risk: how genetic liability is represented and the context in which that liability is expressed. Across multiple biobanks and ancestries, factor-based PGS captured phenomically coherent and portable dimensions of genetic liability, but their predictive value was not universal. Instead, the benefit of phenomic aggregation was disease dependent: it was most evident for asthma, where a respiratory factor PGS better captured shared liability across heterogeneous respiratory phenotypes than a matched disease-specific score, but limited for CAD and T2D, where disease-specific GWAS provided a more complete representation of inherited risk. These findings position phenome-derived PGS not as a general replacement for disease-specific scores, but as a conditional alternative that is most useful when diagnosis-based GWAS incompletely capture underlying liability, particularly in settings where disease definitions are heterogeneous, noisy, or sensitive to ascertainment.

Our framework also differs from existing cross-trait approaches that seek to improve prediction for a prespecified target outcome. Methods such as MTAG^5^ leverage cross-trait genetic correlation at the GWAS stage to improve trait-specific association estimates, whereas multi-trait PRS approaches, such as PRSmix+^3^ and GPS_Mult_^16,17^, combine multiple trait-specific scores at the prediction stage to optimize disease-specific performance. By contrast, we redefine the unit of genetic signal itself by deriving PGS from unsupervised latent phenomic factors and then testing whether these representations better capture disease liability than diagnosis-based GWAS. This distinction matters because our central conclusion is that the value of broader phenomic aggregation varies across diseases.

Asthma provides a clear example of such a setting. The respiratory factor PGS outperformed the matched internal asthma PGS and showed greater cross-ancestry portability, despite substantially smaller discovery samples than the largest available external asthma GWAS^14,18^. This advantage was accompanied by higher SNP-based heritability and broader pathway enrichment, consistent with the possibility that factor-derived GWAS captured syndrome-level liability spread across correlated respiratory and allergic manifestations rather than a single case definition. In this context, improved phenotype definition and stronger GWAS signal may have outweighed the expected cross-trait and cross-ancestry prediction disadvantage imposed by incomplete genetic correlation with asthma across populations^19,20^. More broadly, these findings suggest that phenotype definition, and not only discovery sample size, is an important determinant of PGS portability.

An additional implication is that portability of factor-derived PGS does not require transferability of the underlying phenotypic factor structure itself. Although latent factors derived from deep phenotyping are cohort specific, the genetic liabilities they capture can still manifest as reproducible phenomic signatures across independent datasets. This decoupling between factor construction and external phenomic manifestation suggests that phenome-derived PGS may provide a compact and biologically interpretable representation of liability even when the original phenotypic architecture cannot be reconstructed outside the discovery cohort^6^. More generally, differences across biobanks likely reflect not only genetic architecture and phenotype definition, but also cohort-specific ascertainment and healthcare-system context^21^.

By contrast, CAD and T2D illustrate settings in which disease-specific GWAS remain more informative than phenomic aggregation. In both diseases, the internally derived disease-specific PGS outperformed the corresponding factor-based score, and larger public GWAS-based PGS further outperformed both. In both cases, the corresponding factors appeared to capture clinically broader mixtures of diagnosis, treatment, and comorbidity burden than the more specific inherited liability represented by disease-specific GWAS. When disease-specific GWAS already provide a strong and relatively complete model of inherited risk, broader phenomic representation offers limited additional value. The concordance between predictive performance, genetic correlation, SNP heritability and pathway enrichment supports this interpretation and helps define the conditions under which phenome-derived PGS are most likely to improve disease risk prediction.

Not all factor-derived PGS, however, are equally interpretable in biological terms. In contrast to disease-oriented factors, which are plausibly anchored in shared pathophysiology, PGS derived from socio-environmental factors may capture a mixture of behavioral predisposition, socially patterned exposures and residual confounding related to population structure^6,22,23^. Genetic associations for socio-behavioral traits, including educational attainment and substance use, are known to reflect a combination of direct genetic effects, gene-environment correlation and environmental stratification^24–26^. Their broad associations, including enrichment for mental and behavioral disorders across biobanks, may therefore reflect both meaningful predictive structure and non-causal pathways. This distinction is important for interpreting phenome-derived PGS more broadly and motivates the need to model social context explicitly when evaluating the practical meaning of genetic risk.

A second major finding, observed in AoU, is that SDoH contributed substantial and largely independent predictive information beyond the disease-optimal genetic model. Across asthma, CAD and T2D, SDoH explained liability-scale variance comparable in magnitude to single PGS and showed only limited overlap with the optimal genetic predictor. The weak correlation between PGS and SDoH, together with their joint predictive contribution, indicates that social context is not simply acting as a proxy for inherited liability, but instead captures an additional and largely orthogonal dimension of disease risk. Within the AoU setting, these results extend prior work on context-dependent PGS performance^7,9^ by demonstrating that structured SDoH measures can be incorporated explicitly as a complementary component of liability rather than treated as unmodeled background heterogeneity.

A particularly important implication is that social context altered not only the incremental predictive value of PGS, but also its calibration. For asthma and CAD, increasing SDoH burden attenuated the incremental AUC contributed by PGS and shifted the relationship between genetic percentile and predicted prevalence. For T2D, this attenuation was weaker, indicating relative preservation of genetic stratification despite increasing social burden. Across all three diseases, however, the same genetic percentile did not correspond to the same predicted prevalence across social strata. This is clinically important because it means that the interpretation of PGS is context dependent even when the score itself is fixed. In this setting, SDoH is not merely an additive predictor, but a determinant of how genetic risk should be interpreted, calibrated and translated into predicted prevalence. More broadly, these findings show that structured social context can systematically reshape the practical meaning of genetic risk, although the magnitude and form of this effect will depend on the population and measurement context in which SDoH are assessed^27^. Accordingly, these estimates should be interpreted as context-dependent rather than assumed to transfer unchanged across healthcare systems or populations.

Importantly, these context-dependent patterns are scale dependent. For asthma and CAD, genetic stratification as measured by incremental AUC and odds-ratio gradients attenuated with increasing SDoH burden, whereas predicted-prevalence differences across genetic strata were preserved despite rising baseline prevalence across social strata. T2D showed a different pattern, with weaker attenuation of relative genetic stratification and a larger increase in predicted-prevalence differences across SDoH strata. These findings are therefore consistent rather than contradictory, and emphasize that clinically relevant effect modification may be more apparent on additive or predicted-prevalence-difference scales than on multiplicative scales alone.

These results have two broader implications. First, they argue against one-size-fits-all approaches to polygenic prediction, consistent with prior recommendations emphasizing trait-specific model optimization^20,28^. The optimal genetic representation of disease risk differs across outcomes, and broader aggregation across phenotypes is most useful when it captures coherent latent liability that is only partially represented by diagnosis-based GWAS. Second, they show that improving polygenic prediction requires attention not only to genetic architecture, but also to the contexts in which predictions are applied and interpreted. Together, these findings suggest that evaluation of PGS should integrate both representation of liability and social context.

Several limitations should be noted. First, comprehensive SDoH measurement in AoU was available primarily in EUR, and cross-ancestry analyses relied on a reduced harmonized subset of variables; accordingly, our cross-population estimates of social-context effects are likely conservative. Second, SDoH data in AoU were assembled across multiple surveys with differing completion rates, leading to substantial and non-uniform missingness across items. Although we used multiple imputation in the primary analyses, missingness related to survey participation may still introduce selection bias or imperfect recovery of the underlying social context. Third, the primary SDoH analyses in AoU were performed under a prevalent-disease framework, so standardized predicted prevalence and calibration patterns should not be interpreted as incident absolute risk over a fixed prediction horizon. Although the temporally robust subset and incident-case sensitivity analyses reduce concern about reverse causation, they do not fully resolve temporal ordering between measured SDoH exposure and disease onset. Fourth, our analyses were designed to evaluate prediction rather than causal mechanisms, and therefore do not identify the biological or social pathways through which inherited liability and social exposures combine to influence disease.

Overall, our results indicate that polygenic prediction is shaped by both the representation of genetic liability and the context in which that liability is expressed. Phenome-derived PGS improve prediction under specific architectural conditions, whereas social context independently modifies the performance, calibration and interpretation of genetic risk, with the strongest evidence here arising from analyses in AoU. Together, these findings support a framework in which the practical meaning and utility of polygenic prediction are jointly determined by genetic architecture and the environments in which risk is realized.

## Methods

### Study design

We evaluated disease prediction along two complementary axes: how genetic liability is represented and the social context in which that liability is expressed. First, we constructed factor-based polygenic scores (PGS) from genome-wide association studies (GWAS) of latent phenomic factors derived in UK Biobank (UKB), together with internal and external disease-specific benchmarking scores. Second, we assessed the phenomic specificity, portability, and predictive performance of factor-based PGS across biobanks and ancestry groups. Third, in the All of Us Research Program (AoU), we integrated social determinants of health (SDoH) with the disease-optimal genetic model to evaluate independent and joint contributions, interaction, calibration, and context-dependent stratification. All analyses were performed within biobank-by-ancestry strata.

### Cohorts and quality controls

#### UK Biobank

UK Biobank (UKB) is a population-based cohort of approximately 500,000 individuals aged 40-69 years at recruitment^29^. Thirty-five statistically independent latent factors were previously derived from 2,772 phenotypes using exploratory and confirmatory factor analysis in 361,144 unrelated individuals of European genetic ancestry^6^. Factor scores were calculated as weighted linear combinations of contributing items.

GWAS of factor scores were performed using weighted least-squares regression implemented in Hail (https://hail.is/), adjusting for age, sex, age², age×sex, age²×sex, assessment center, and the first 20 genetic principal components (PCs). The factor-analysis cohort served as the discovery dataset for PGS construction. Independent target sets were defined by excluding related individuals and applying the same PCA-based ancestry selection criteria detailed elsewhere (https://github.com/Nealelab/UK_Biobank_GWAS/blob/master/ukb31063_eur_selection.R), yielding 8,718 European ancestry (EUR) individuals and additional non-European ancestry groups: 6,258 African (AFR), 988 Admixed American (AMR), 8,239 Central/South Asian (CSA), 2,683 East Asian (EAS), and 1,565 Middle Eastern (MID).

#### All of Us Research Program

The All of Us (AoU) Research Program integrates survey, physical measurement, and electronic health record (EHR) data^30,31^. Among 245,350 individuals with whole-genome sequencing data (April 2023 release v7), genetic ancestry was assigned using projection onto reference PCs derived from 1000 Genomes Phase 3 and Hume Genome Diversity Project panels, followed by random forest classification and within-cluster refinement among unrelated individuals (kinship coefficient <0.1) as described in the Pan-UK Biobank project^32^. Final ancestry-stratified sample sizes were 112,634 EUR, 44,436 AFR, 35,426 AMR, 2,056 CSA, 5,009 EAS, and 410 MID.

#### Mass General Brigham Biobank

The Mass General Brigham Biobank (MGBB) constitutes a hospital-based biorepository spanning the Mass General Brigham healthcare network in the greater Boston area^33–35^. We analyzed 53,281 individuals genotyped on the Illumina GSA array. Genetic ancestry assignment followed the same projection-based framework used in AoU, and related individuals were excluded (KING kinship coefficient >0.084). Final ancestry groups included 37,061 EUR, 2,252 AFR, 2,601 AMR, 514 CSA, 999 EAS, and 239 MID.

#### Genotype quality control

Variants were retained if they satisfied minor allele frequency >0.01, Hardy-Weinberg equilibrium P>1×10⁻⁶, genotype missing rate <0.05, and imputation quality thresholds (INFO>0.8 in UKB; INFO>0.3 in MGBB). Analyses were restricted to HapMap3 variants. Individuals with genotype missingness >0.1 were excluded. Alleles were harmonized across summary statistics and target genotypes; ambiguous SNPs were removed when necessary.

#### Phenotype definitions

Disease outcomes were defined using PheCodes mapped from ICD-9 and ICD-10 diagnoses via the PheWAS Catalog (https://phewascatalog.org/). Individuals were classified as cases if ≥1 corresponding ICD code was present. Within each biobank-by-ancestry stratum, PheCodes with ≥50 cases were retained for analysis. UKB PheCodes were obtained from the pan-UKB pipeline (https://github.com/atgu/ukbb_pan_ancestry/blob/master/assign_PheCodes.py). The numbers of evaluated PheCodes were 1,232 in UKB, 3,293 in MGBB, and 5,450 in AoU, reflecting differences in sample size and clinical ascertainment across biobanks.

Unless otherwise noted, primary disease prediction, calibration, and risk-stratification analyses were based on prevalent case definitions. Incident-case analyses were performed separately using time-to-event survival models, as described below.

### Construction of polygenic scores

#### Factor-based PGS

Factor-based PGS were constructed from UKB factor GWAS summary statistics using PRS-CS (auto model) with the provided European LD reference panel^13^. Default parameters were used, and the global shrinkage parameter was inferred from the data. Scores were generated in independent target datasets across ancestries per biobank and standardized within ancestry strata using control distributions.

#### Disease-specific PGS

For benchmarking, internal disease-specific GWAS were performed for asthma, coronary artery disease (CAD), and type 2 diabetes (T2D) within UKB using identical QC, ancestry assignment, and covariate adjustments as factor GWAS. External disease-specific PGS were constructed using large-scale published GWAS summary statistics (**Table S4**)^18,36,37^. All PGS were standardized within ancestry strata prior to downstream analyses.

### Phenome-wide association and enrichment analyses

Within each biobank-by-ancestry stratum, associations between factor-based PGS and PheCodes were tested using logistic regression adjusting for age, sex, quadratic age terms, age-by-sex interactions (where applicable), and the first 10 PCs. Effect sizes are reported as odds ratios (OR) per standard deviation increase in PGS with 95% confidence intervals. Statistical significance was assessed using Bonferroni correction within each stratum. To visualize these associations, we employed the PheWAS R package^38^ to generate Manhattan plots.

To characterize phenomic structure, enrichment of significant associations across PheCode chapters was quantified within each stratum and combined across biobanks within ancestry using Stouffer’s method. Full enrichment formulas and meta-analytic procedures are provided in **Supplementary Note 1**.

### Candidate outcomes and predictive evaluation

Because factor-based PGS were derived without reference to prespecified disease endpoints, we first identified candidate anchor outcomes empirically on the basis of Bonferroni-significant PGS associations, replication in at least two biobanks, and concordance with dominant factor loadings, defined as agreement between the highest-loading phenotypes within a factor and its strongest disease associations. We then selected asthma, coronary artery disease (CAD), and type 2 diabetes (T2D) as the primary outcomes for comparative benchmarking because they represented clinically important diseases across distinct domains and spanned settings in which factor-based prediction was advantageous as well as settings in which disease-specific prediction remained superior. Additional qualifying factor-outcome pairs were treated as supporting examples and are presented in the Extended Data and Supplementary Material.

Within each biobank-by-ancestry stratum, logistic regression models adjusted for age, sex, quadratic age terms, age-by-sex interactions, and the first 10 PCs were used to evaluate predictive performance of factor-based PGS, disease-specific PGS, and joint models including both scores. Model performance was assessed using Nagelkerke’s pseudo-*R*^2^, *R*^2^ on the liability scale^39^, area under the receiver operating characteristic curve (AUC), and odds ratios per standard deviation increase in PGS. Incremental performance was defined relative to the covariate-only model. For liability-scale transformations, the proportion of cases in the individual population in each biobank was used to approximate disease prevalence. Confidence intervals were estimated via nonparametric bootstrap resampling. Likelihood ratio tests were used to assess incremental predictive value when adding one PGS to models containing the other. Correlation between factor-based and disease-specific PGS was evaluated using Pearson correlation coefficients within ancestry strata. To assess portability, predictive performance was compared across biobanks and ancestry groups. Relative performance retention was quantified as the ratio of *R*^2^ on the liability scale in non-European ancestry groups to that observed in European ancestry samples within the same biobank.

### SNP heritability and genetic correlation

To characterize genetic architecture differences underlying performance patterns between factor and disease-specific PGS, we estimated SNP-based heritability (ℎ^2^) and genetic correlation (*r*_g_) using LD score regression (LDSC v1.0.1)^40,41^. The 1000 Genomes Phase 3 European reference panel was used as the reference panel. SNP heritability was estimated on the observed scale for quantitative traits and transformed to the liability scale for binary traits using cohort-level prevalence estimates. Genetic correlation between factor and disease GWAS was estimated using bi-variate LDSC.

### MAGMA gene and pathway enrichment

Gene-level association and competitive gene-set enrichment analyses were performed using MAGMA (v1.10)^42^ for both factor GWAS and internal disease GWAS summary statistics. Multiple testing was controlled using Benjamini-Hochberg false discovery rate correction (FDR<0.05)^43^.

Identical workflows were applied to enable direct comparison between factor-based and disease-specific genetic architecture.

### Social determinants of health

Social determinants of health (SDoH) were evaluated in the All of Us Research Program (AoU), where structured social measures were available at sufficient depth for joint modeling with genetic risk. SDoH were operationalized using the Healthy People 2030 framework (HP2030) spanning five domains: Economic Stability, Education Access and Quality, Healthcare Access and Quality, Neighborhood and Built Environment, and Social and Community Context^44^. Candidate SDoH variables were initially reviewed across five All of Us surveys (The Basics, Lifestyle, Overall Health, Healthcare Access & Utilization, and Social Determinants of Health; release v7; (https://www.researchallofus.org/data-tools/survey-explorer/) by referencing the systematic mapping developed by Hysong et al.^45^, which aligned All of Us survey items to HP2030 objectives. The final analytic SDoH set was drawn from three of these surveys, as detailed in **Table S6**.

Primary European ancestry analyses used 29 SDoH variables chosen to provide broad coverage across all five HP2030 domains. A temporally robust 16-variable subset was selected for sensitivity analyses designed to reduce, though not eliminate, concern about reverse causation in the prevalent-disease analyses. Those variables were selected on the basis of questionnaire content and reference frame, prioritizing indicators intended to reflect relatively stable structural or socioeconomic conditions and excluding items more likely to capture short-term states or consequences of current illness. This classification was based on item wording and conceptual stability rather than confirmed temporal ordering relative to disease onset. Because the primary analyses use prevalent disease, even this restricted subset does not establish that measured SDoH exposure preceded disease onset; instead, it serves as a sensitivity analysis designed to reduce susceptibility to reverse causation. Cross-ancestry analyses used a harmonized 7-variable subset from the AoU Basics survey with <1% missingness across populations. Variables were recoded so higher values indicated greater social adversity.

For the 29-variable and 16-variable analyses, missing values were addressed using multiple imputation by chained equations (M = 50 imputations). The 7-variable cross-ancestry analysis was conducted using complete-case data because missingness was minimal. We first created domain-specific composite scores by averaging standardized values of all variables within each of the five HP2030 domains. An overall SDoH composite score was then constructed by averaging the five domain scores, calculated only for participants with valid scores on all five domains. This approach gives equal weight to each domain regardless of the number of indicators per domain, consistent with the HP2030 framework’s conceptualization of SDoH domains as multidimensional but equally important contributors to social burden.

Detailed procedures for variable selection, coding, harmonization, imputation, control-based re-standardization, and benchmarking of the 16-variable and 7-variable composites relative to the full 29-variable representation are provided in **Supplementary Notes 2-5**.

### Joint PGS and SDoH modeling

Joint analyses of genetic and social risk were performed in AoU within biobank-by-ancestry strata. For these analyses, the disease-optimal PGS was defined as the factor-based PGS for asthma and the disease-specific PGS for coronary artery disease (CAD) and type 2 diabetes (T2D), on the basis of the internal benchmarking analyses described above. Unless otherwise noted, these analyses were conducted under a prevalent-disease framework. We fit nested logistic regression models including baseline covariates, the disease-optimal PGS, the SDoH composite, and their multiplicative interaction term (PGS×SDoH). For imputed analyses, results were pooled across imputations using Rubin’s rules^46^. We quantified predictive performance using area under the receiver operating characteristic curve (AUC), liability-scale *R*^2^, and regression coefficients or odds ratios as appropriate.

To assess whether social context modified the predictive meaning of genetic liability, we evaluated four complementary features of joint genetic-social architecture: (i) the independent contribution of SDoH beyond the disease-optimal PGS; (ii) PGS×SDoH interaction effects; (iii) stratified predictive performance and odds-ratio gradients across tertiles of SDoH burden; and (iv) calibration and standardized predicted prevalence across joint strata of PGS decile and SDoH tertile. We additionally examined high-risk tails of the PGS and joint-prediction distributions.

To characterize the joint relationship between genetic and social risk on the prevalence scale, we estimated standardized predicted prevalence within each joint stratum of PGS decile *g* and SDoH tertile *s* by averaging fitted probabilities from the logistic model over the empirical covariate distribution of the analysis sample:

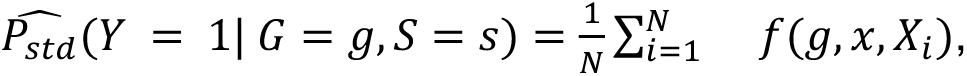

where *f*(⋅) denotes the fitted probability from the regression model, *X*_*i*_ denotes the covariates for individual *i*, and *N* is the total number of individuals in the analysis sample. Thus, for each joint stratum, PGS decile and SDoH tertile were fixed while age, sex, and other adjustment covariates were standardized to the observed covariate distribution in the full sample. We refer to these quantities as standardized predicted prevalence rather than absolute risk because the primary analyses are based on prevalent disease status rather than incident disease over a fixed prediction horizon.

As a prospective sensitivity analysis, we separately fit incident-case Cox proportional hazards models in AoU-EUR after excluding prevalent cases at baseline. These analyses were used to evaluate whether the principal genetic-social patterns observed in the prevalent-disease analyses extended to future disease risk.

Detailed implementation of interaction testing, stratified performance analyses, standardized predicted-prevalence grids, calibration metrics, high-risk tail analyses, and incident-case Cox models is described in **Supplementary Notes 6-11**.

## Data availability

Individual-level genotype and phenotype data from UK Biobank are available to approved researchers through application to the UK Biobank (Application no. 31063) at https://www.ukbiobank.ac.uk, subject to the UK Biobank data access policies. All of Us Research Program data are available to registered researchers through the All of Us Researcher Workbench at https://www.researchallofus.org, in accordance with the program’s data use agreement. Mass General Brigham Biobank data are available upon application to the institutional data access committee and subject to institutional review and data use policies. External GWAS summary statistics used for disease-specific polygenic score construction (asthma, coronary artery disease, and type 2 diabetes) are publicly available from the original publications and repositories listed in **Table S4**. Source data underlying the main figures and supplementary tables generated in this study, together with code for polygenic score construction, joint PGS-SDoH modeling, stratified performance analyses, calibration assessment, and standardized predicted-prevalence analyses, will be made publicly available at Zenodo upon publication.

## Acknowledgements

Y.W. is supported in part by the NIH-NHGRI K99HG013969. A.R.M is supported by the K99/R00MH117229. K.T. is supported by F31HL167378 and by the ECOR Claflin Award to A.R.M. A.R.M. and Y.W. are supported by U01HG011719. Additional support for this work to A.R.M. and Y.W. also comes from the European Union’s Horizon 2020 research and innovation program under grant agreement 101016775 (INTERVENE Consortium). This research was supported in part by the Intramural Research Program of the National Institutes of Health (NIH). The contributions of the NIH authors are considered Works of the United States Government. The findings and conclusions presented in this paper are those of the author and do not necessarily reflect the views of the NIH or the U.S. Department of Health and Human Services. We thank the participants and investigators of the UK Biobank, Mass General Brigham Biobank, and the All of Us Research Program for enabling this research.

## References

1. Forrest, I. S. et al. Machine learning-based marker for coronary artery disease: derivation and validation in two longitudinal cohorts. Lancet 0, (2022).

2. Lee, C. H. et al. Liability threshold model-based disease risk prediction based on electronic health record phenotypes. Nat. Genet. 57, 2872–2881 (2025).

3. Truong, B. et al. Integrative polygenic risk score improves the prediction accuracy of complex traits and diseases. Cell Genom. 4, 100523 (2024).

4. Krapohl, E. et al. Multi-polygenic score approach to trait prediction. Mol. Psychiatry 23, 1368–1374 (2017).

5. Turley, P. et al. Multi-trait analysis of genome-wide association summary statistics using MTAG. Nat. Genet. 50, 229–237 (2018).

6. Carey, C. E. et al. Principled distillation of UK Biobank phenotype data reveals underlying structure in human variation. Nat. Hum. Behav. 8, 1599–1615 (2024).

7. Mostafavi, H. et al. Variable prediction accuracy of polygenic scores within an ancestry group. Elife 9, e48376 (2020).

8. Hui, D. et al. Quantifying factors that affect polygenic risk score performance across diverse ancestries and age groups for body mass index. Pac. Symp. Biocomput. 28, 437–448 (2023).

9. Hou, K. et al. Calibrated prediction intervals for polygenic scores across diverse contexts. Nat. Genet. 56, 1386–1396 (2024).

10. Gómez, C. A. et al. Addressing health equity and social determinants of health through Healthy People 2030. J. Public Health Manag. Pract. 27, S249–S257 (2021).

11. Inouye, M. et al. Genomic Risk Prediction of Coronary Artery Disease in 480,000 Adults: Implications for Primary Prevention. J. Am. Coll. Cardiol. 72, 1883–1893 (2018).

12. He, Y. et al. Comparisons of Polyexposure, Polygenic, and Clinical Risk Scores in Risk Prediction of Type 2 Diabetes. Diabetes Care 44, 935–943 (2021).

13. Ge, T., Chen, C.-Y., Ni, Y., Feng, Y.-C. A. & Smoller, J. W. Polygenic prediction via Bayesian regression and continuous shrinkage priors. Nat. Commun. 10, 1776 (2019).

14. Tsuo, K. et al. Multi-ancestry meta-analysis of asthma identifies novel associations and highlights the value of increased power and diversity. Cell Genomics 2, 100212 (2022).

15. Wang, Y. et al. Global Biobank analyses provide lessons for developing polygenic risk scores across diverse cohorts. Cell Genomics 3, 100241 (2023).

16. Patel, A. P. et al. A multi-ancestry polygenic risk score improves risk prediction for coronary artery disease. Nat. Med. 29, 1793–1803 (2023).

17. Zhang, Z. Variable selection with stepwise and best subset approaches. Ann. Transl. Med. 4, 136 (2016).

18. Zhou, W. et al. Global Biobank Meta-analysis Initiative: Powering genetic discovery across human disease. Cell Genom 2, 100192 (2022).

19. Wang, Y., Tsuo, K., Kanai, M., Neale, B. M. & Martin, A. R. Challenges and Opportunities for Developing More Generalizable Polygenic Risk Scores. Annu Rev Biomed Data Sci 5, 293–320 (2022).

20. Wang, Y. et al. Polygenic prediction across populations is influenced by ancestry, genetic architecture, and methodology. Cell Genomics 100408 (2023) doi:10.1016/j.xgen.2023.100408.

21. Pimplaskar, A. et al. Inclusion bias affects common variant discovery and replication in a health-system linked biobank. Am. J. Hum. Genet. 113, 702–714 (2026).

22. Berg, J. J. et al. Reduced signal for polygenic adaptation of height in UK Biobank. Elife 8, (2019).

23. Sohail, M. et al. Polygenic adaptation on height is overestimated due to uncorrected stratification in genome-wide association studies. Elife 8, (2019).

24. Lee, J. J. et al. Gene discovery and polygenic prediction from a genome-wide association study of educational attainment in 1.1 million individuals. Nat. Genet. 50, 1112–1121 (2018).

25. Abdellaoui, A. et al. Genetic correlates of social stratification in Great Britain. Nat. Hum. Behav. 3, 1332–1342 (2019).

26. Young, A. I., Benonisdottir, S., Przeworski, M. & Kong, A. Deconstructing the sources of genotype-phenotype associations in humans. Science 365, 1396–1400 (2019).

27. Namba, S. et al. A cross-population compendium of gene-environment interactions. Nature 651, 688–697 (2026).

28. Tsuo, K., et al. All of Us diversity and scale improve polygenic prediction contextually with greatest improvements for under-represented populations. bioRxivorg (2024) doi:10.1101/2024.08.06.606846.

29. Bycroft, C. et al. The UK Biobank resource with deep phenotyping and genomic data. Nature 562, 203–209 (2018).

30. Genomic data in the All of Us Research Program. Nature 1–7 (2024) doi:10.1038/s41586-023-06957-x.

31. All of Us Research Program Investigators et al. The ‘All of Us’ Research Program. N. Engl. J. Med. 381, 668–676 (2019).

32. Karczewski, K. J. et al. Pan-UK Biobank genome-wide association analyses enhance discovery and resolution of ancestry-enriched effects. Nat. Genet. 57, 2408–2417 (2025).

33. Castro, V. M. et al. The Mass General Brigham Biobank Portal: an i2b2-based data repository linking disparate and high-dimensional patient data to support multimodal analytics. J. Am. Med. Inform. Assoc. 29, 643–651 (2022).

34. Boutin, N. T. et al. The Evolution of a Large Biobank at Mass General Brigham. J Pers Med 12, (2022).

35. Koyama, S. et al. Genetics and context for precision health in Greater Boston. Nat. Commun. 16, 11661 (2025).

36. Aragam, K. G. et al. Discovery and systematic characterization of risk variants and genes for coronary artery disease in over a million participants. Nat. Genet. 54, 1803–1815 (2022).

37. Mahajan, A. et al. Multi-ancestry genetic study of type 2 diabetes highlights the power of diverse populations for discovery and translation. Nat. Genet. 54, 560–572 (2022).

38. Carroll, R. J., Bastarache, L. & Denny, J. C. R PheWAS: data analysis and plotting tools for phenome-wide association studies in the R environment. Bioinformatics 30, 2375–2376 (2014).

39. Lee, S. H., Goddard, M. E., Wray, N. R. & Visscher, P. M. A better coefficient of determination for genetic profile analysis. Genet. Epidemiol. 36, 214–224 (2012).

40. Bulik-Sullivan, B. K. et al. LD Score regression distinguishes confounding from polygenicity in genome-wide association studies. Nat. Genet. 47, 291–295 (2015).

41. Bulik-Sullivan, B. et al. An atlas of genetic correlations across human diseases and traits. Nat. Genet. 47, 1236–1241 (2015).

42. de Leeuw, C. A., Mooij, J. M., Heskes, T. & Posthuma, D. MAGMA: generalized gene-set analysis of GWAS data. PLoS Comput. Biol. 11, e1004219 (2015).

43. Benjamini, Y. & Hochberg, Y. Controlling the false discovery rate: A practical and powerful approach to multiple testing. J. R. Stat. Soc. Series B Stat. Methodol. 57, 289–300 (1995).

44. Social Determinants of Health. https://odphp.health.gov/healthypeople/priority-areas/social-determinants-health.

45. Hysong, M. R. et al. Quantifying social determinants of health for disease prediction: A multi-level approach using Healthy People 2030 and All of us data. medRxiv (2025) doi:10.1101/2025.09.02.25334565.

46. Rubin, D. B. Multiple Imputation for Nonresponse in Surveys. (John Wiley & Sons, Nashville, TN, 1987). doi:10.1002/9780470316696.

