## Extended Data Figures for "Phenome-derived polygenic scores and social determinants jointly shape context-dependent disease risk"

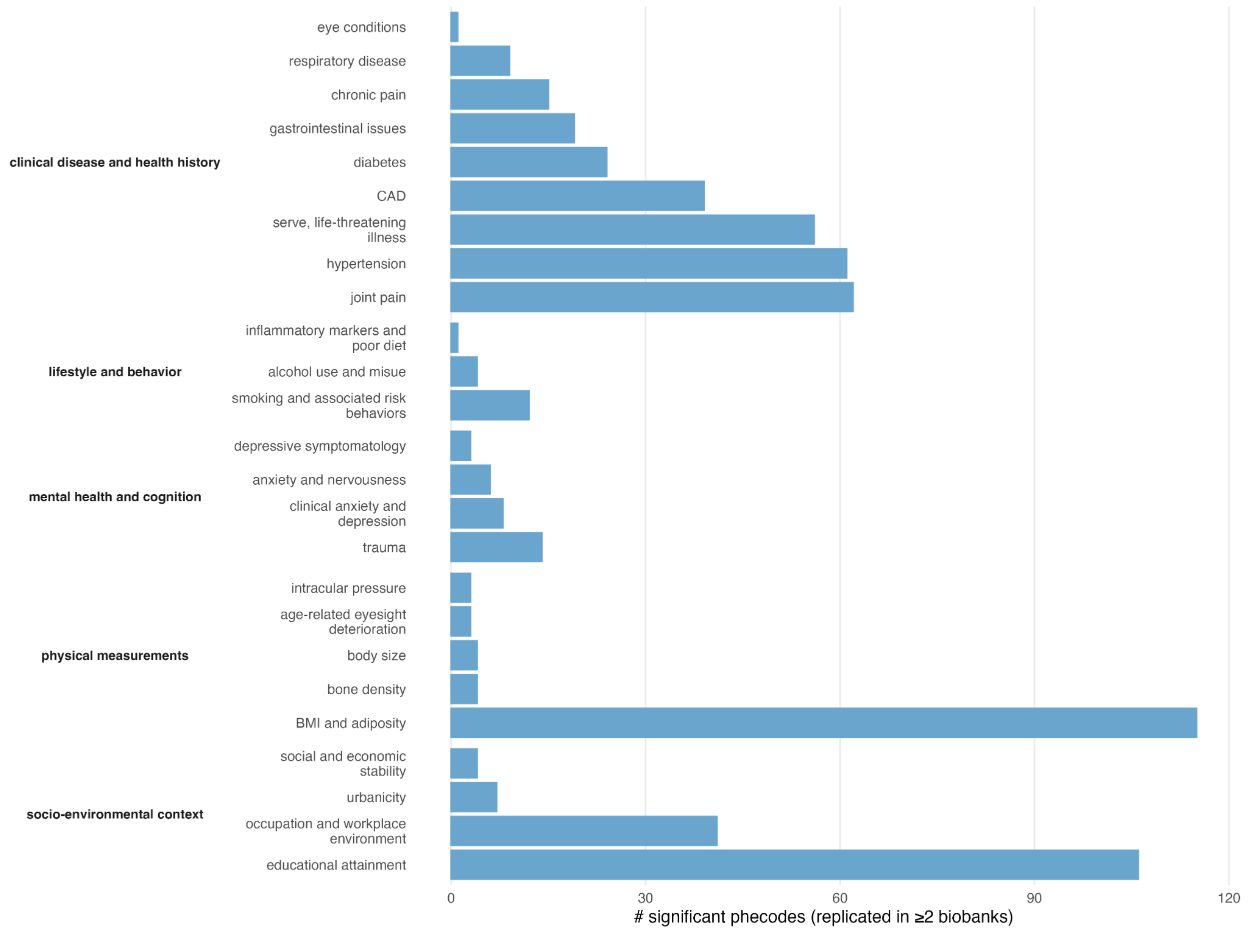


**Extended Data Fig. 1: The counts of significant PGS-phecode associations in individuals of European ancestry**

Bar plot shows, for each latent phenomic factor, the number of phecodes significantly associated with the corresponding factor-based polygenic score in individuals of European ancestry. Factors are grouped by six broad phenomic domains, and bars represent the number of distinct phecodes exhibiting significant associations per factor. The factor within each domain is ranked by the count.

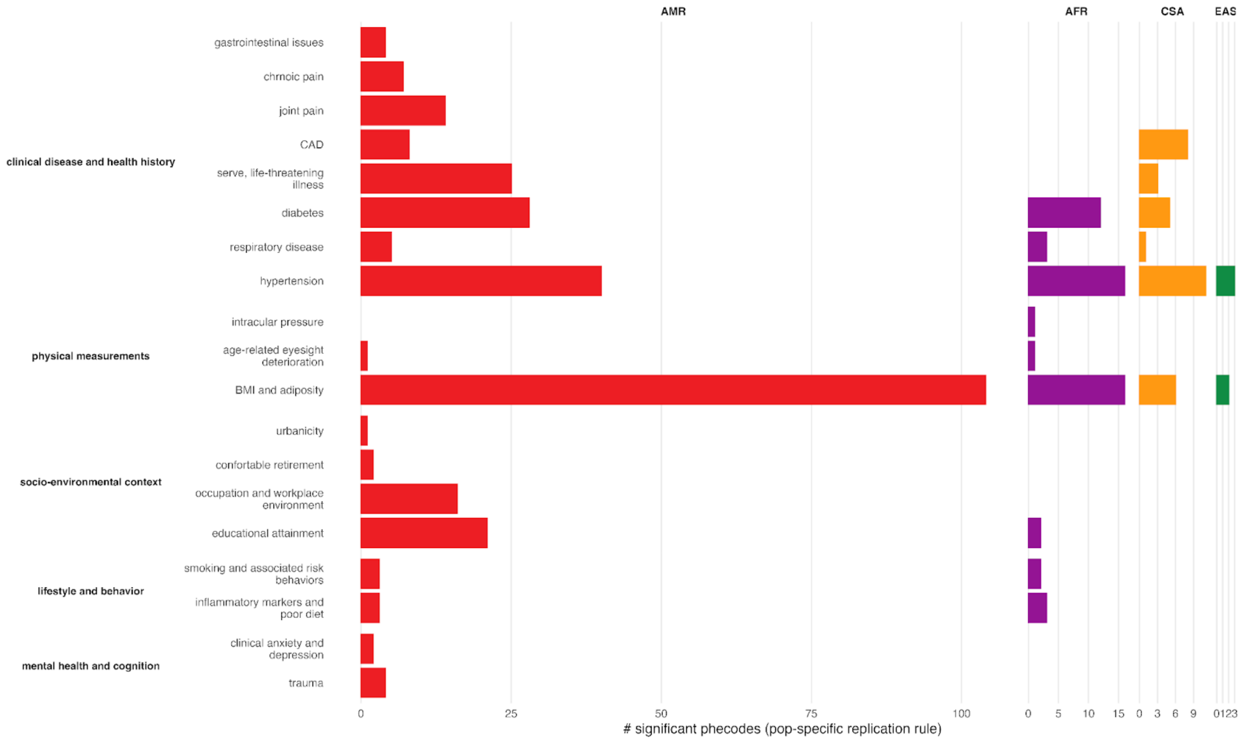


**Extended Data Fig. 2: The counts of significant PGS-phecode associations in individuals of non-European ancestry**

Bar plot shows, for each latent phenomic factor, the number of phecodes significantly associated with the corresponding factor-based polygenic score in individuals of non-European ancestry. Factors are grouped by six broad phenomic domains, and bars represent the number of distinct phecodes exhibiting significant associations per factor. The factor within each domain is ranked by the count within European ancestry.

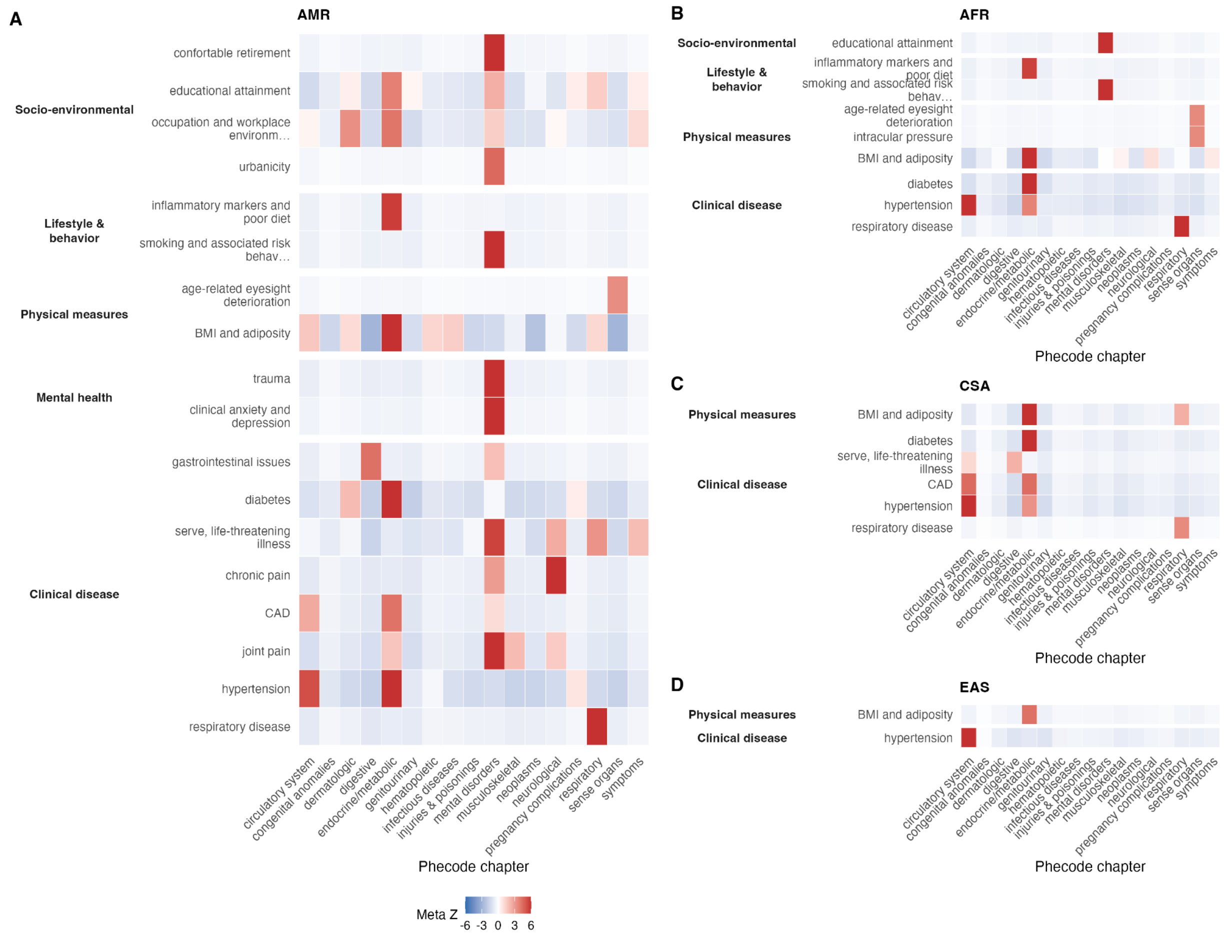


**Extended Data Fig. 3: Meta-analysis of phenomic enrichment in non-European ancestry reveals structured and interpretable genetic liability across disease domains.**

Heatmap shows meta-analytic enrichment Z-scores summarizing the concentration of significant factor-based PGS associations within phecode chapters across biobanks in individuals of non-European ancestry: **A)** Admixed American ancestry, AMR; **B)** African Ancestry, AFR; **C)** Central and South Asian Ancestry, CSA; and **D)** East Asian Ancestry, EAS. Columns correspond to latent phenomic factors grouped into six broad domains, and rows correspond to phecode chapters. Enrichment statistics were meta-analyzed across biobanks using Stouffer's method, and only significant associations identified in at least one biobanks are shown. Color indicates the direction and magnitude of enrichment (Meta Z).

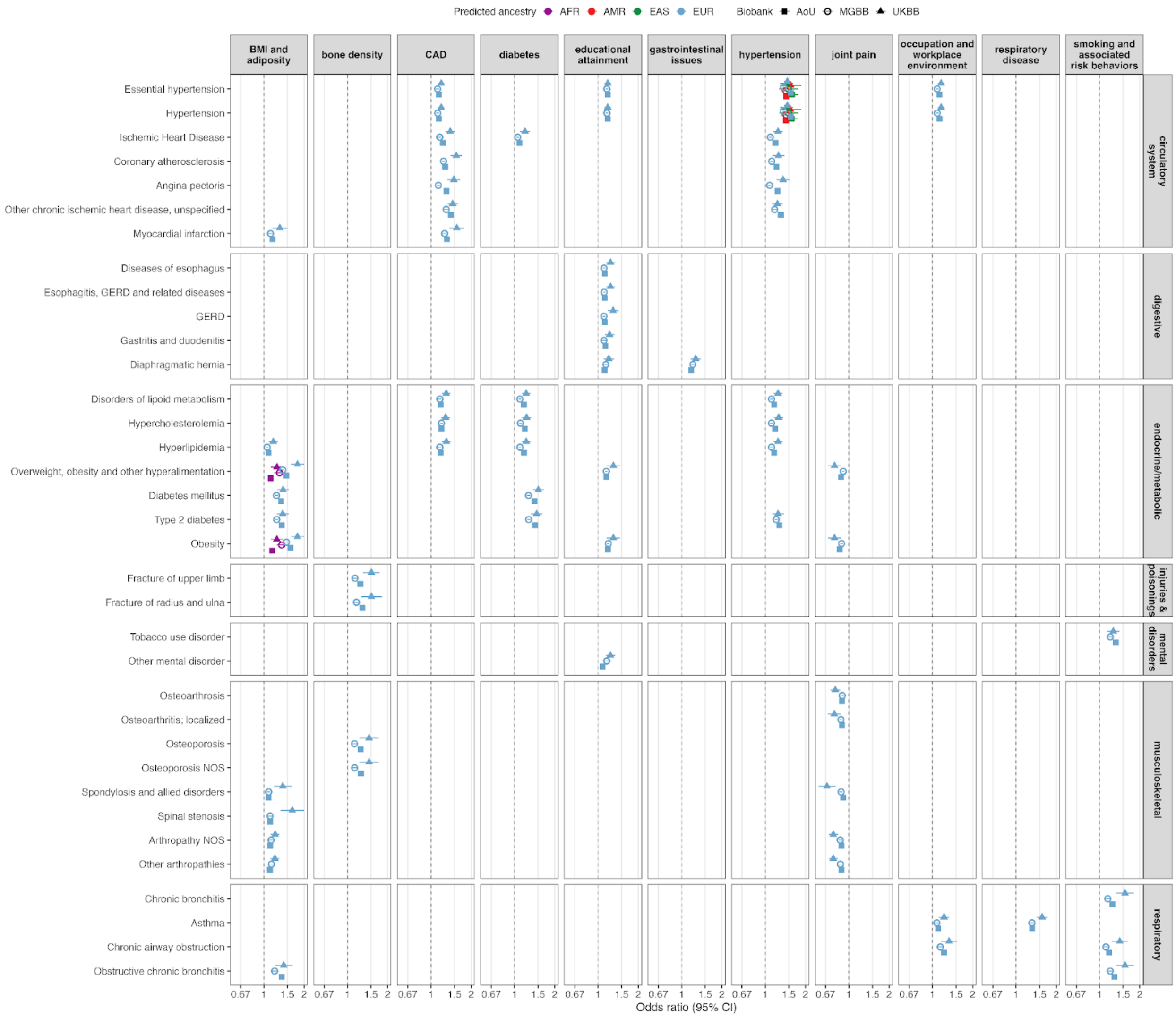


**Extended Data Fig. 4: Shared Significant Associations of PGS-PheCode Across Three Biobanks.**

This figure illustrates the significant correlations between factor-based PGS and specific phecodes identified across all three biobanks. Each column corresponds to a particular factor-based PGS, annotated to describe its factor context. Rows represent individual phecodes within their respective categories. The dashed lines indicate an odds ratio (OR) of 1, serving as a reference point.

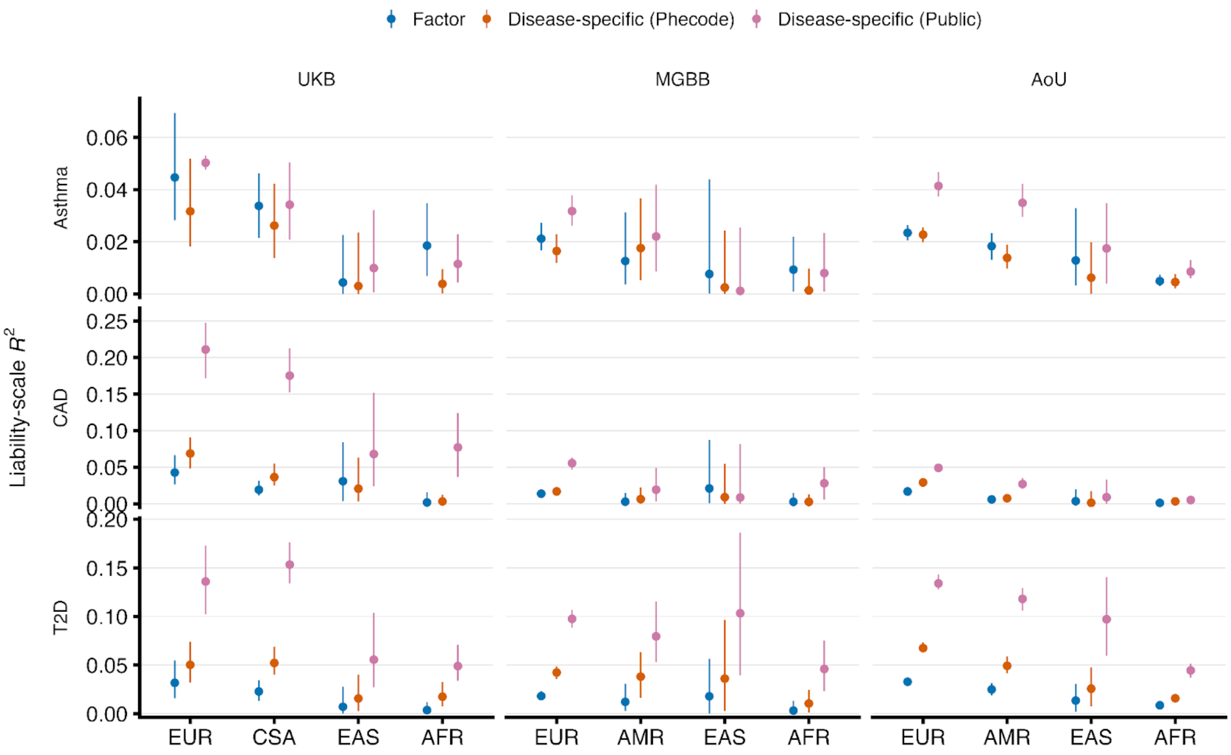


**Extended Data Fig. 5: Cross-biobank performance of factor-based versus disease-specific polygenic scores.**

Liability-scale variance explained for asthma, coronary artery disease (CAD), and type 2 diabetes (T2D) is shown across UK Biobank (UKB), Mass General Brigham Biobank (MGBB), and All of Us (AoU), stratified by genetic ancestry group. Error bars indicate 95% confidence intervals. Blue, factor-based scores derived from latent phenotype factors; orange, disease-specific scores trained using cohort-derived phecodes; pink, disease-specific scores constructed from published GWAS.

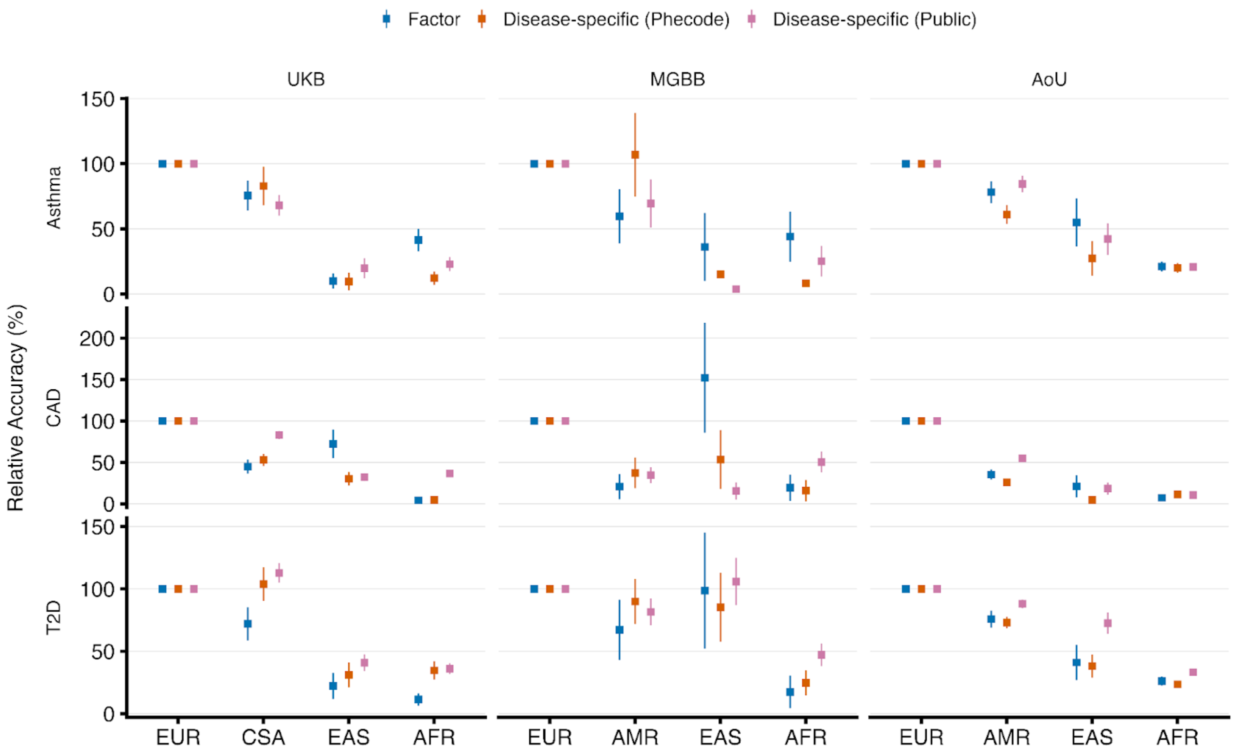


**Extended Data Fig. 6: Relative performance of factor-based and disease-specific polygenic scores across biobanks and ancestries.**

Relative accuracy (%) for asthma, coronary artery disease (CAD), and type 2 diabetes (T2D) is shown in UK Biobank (UKB), Mass General Brigham Biobank (MGBB), and All of Us (AoU), stratified by genetic ancestry group. Relative accuracy is computed as the ratio of $R^2_{liability}$ within each biobank per trait, such that EUR is fixed at 100% and values in other ancestry groups reflect attenuation or preservation of predictive performance relative to EUR. Points denote point estimates and error bars indicate standard errors. Blue, factor-based scores derived from latent phenotype factors; orange, disease-specific scores trained using cohort-derived phecodes; pink, disease-specific scores constructed from published GWAS.

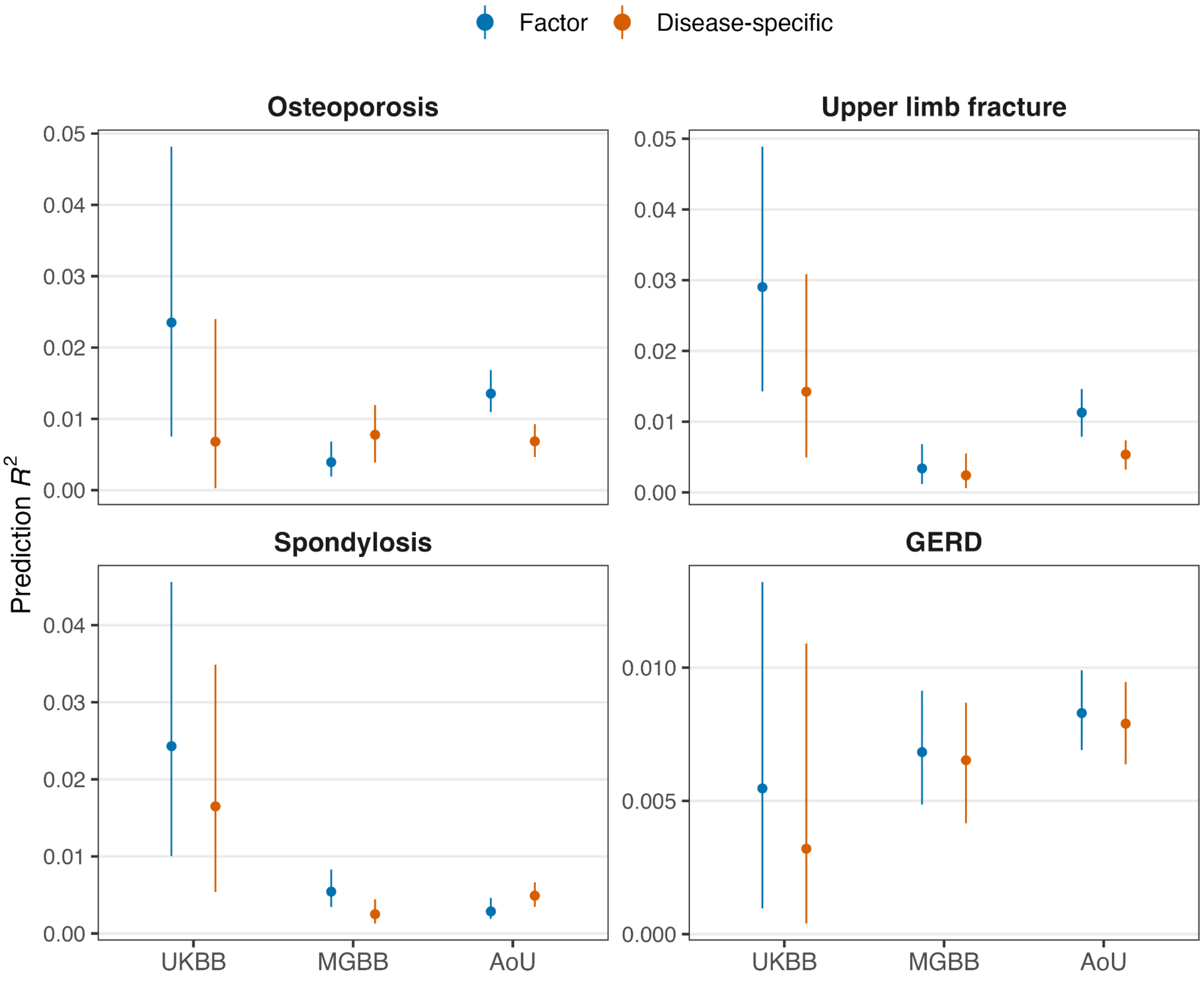


**Extended Data Fig. 7: Additional examples of factor-based versus disease-specific polygenic score performance in European ancestry individuals.**
Liability-scale incremental variance explained for factor-based and disease-specific polygenic scores across biobanks for representative supporting outcomes spanning skeletal, musculoskeletal and gastrointestinal domains. Points indicate point estimates and error bars denote 95% confidence intervals. These examples generally show smaller effect sizes and/or less consistent replication across cohorts than the primary asthma example shown in the main text.

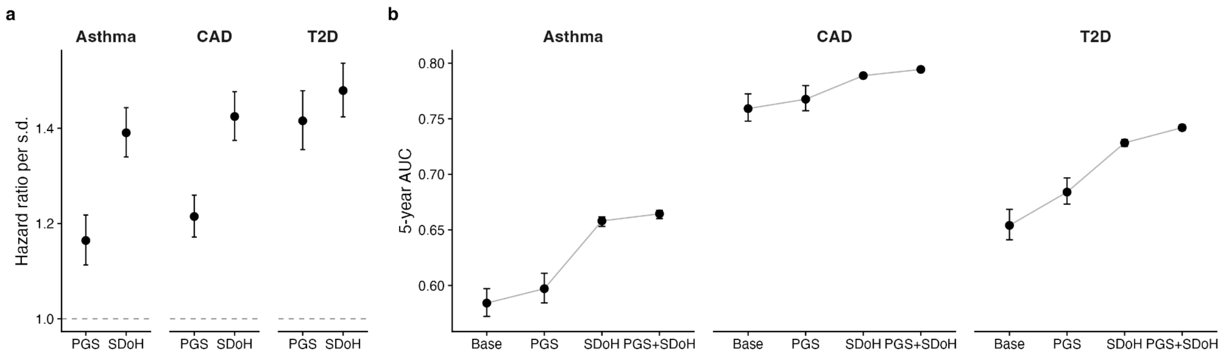


**Extended Data Fig. 8: Incident-disease Cox analyses in AoU-EUR support independent contributions of social and genetic risk.**

**a,** Hazard ratios per standard deviation of the optimal PGS and SDoH from joint Cox models including covariates, the optimal PGS and SDoH. Points indicate estimates and error bars denote 95% confidence intervals. **b,** Five-year AUC for covariate-only, optimal PGS-only, SDoH-only and joint optimal PGS + SDoH models for asthma, CAD and T2D. The optimal PGS was defined from the primary analyses as the factor-derived score for asthma and the disease-specific score for CAD and T2D.

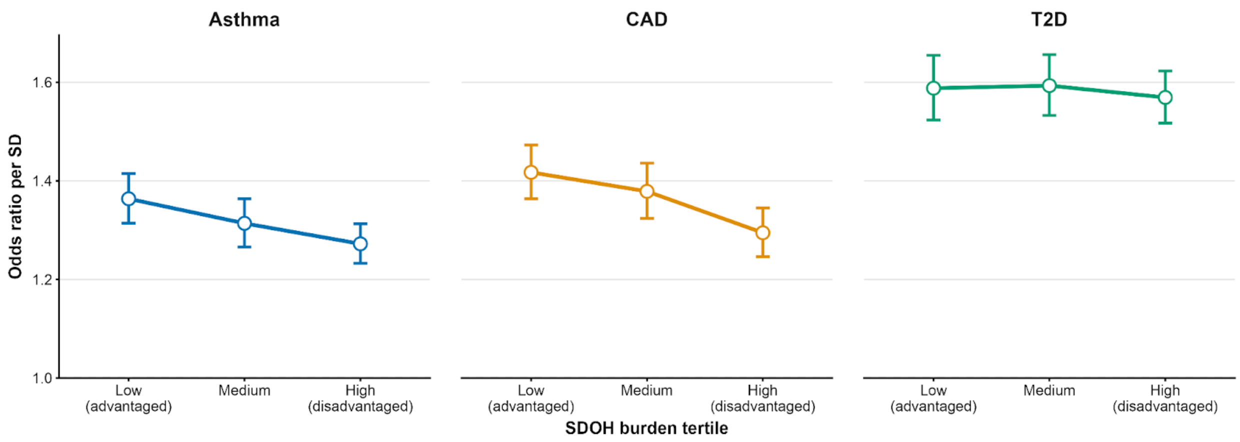


**Extended Data Fig. 9: Social determinants of health modify the genetic effect on disease risk.**

Association between polygenic score (PGS) and disease risk stratified by tertiles of social determinants of health (SDoH) burden for asthma, coronary artery disease (CAD) and type 2 diabetes (T2D). Points indicate odds ratios (ORs) per standard deviation increase in the optimal PGS estimated from logistic regression models within each SDoH tertile (low = most advantaged; high = most disadvantaged), adjusted for age, sex and principal components of ancestry. Error bars denote 95% confidence intervals.

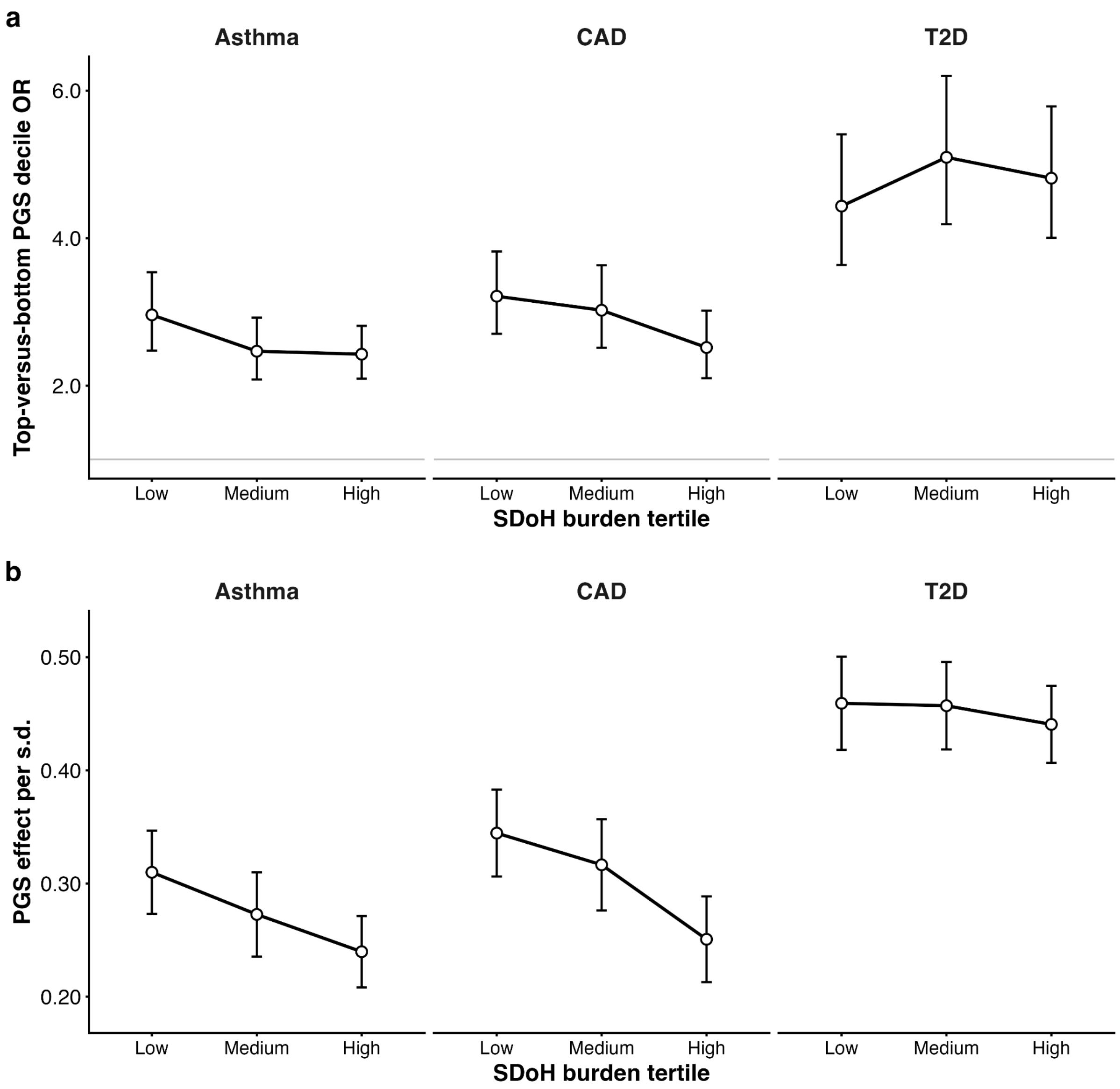


**Extended Data Fig. 10: Stratified genetic effects across social determinants of health burden.**

**a,** Odds ratios comparing individuals in the highest versus lowest decile of the disease-optimal polygenic score (PGS) across tertiles of social determinants of health (SDoH) burden for asthma, coronary artery disease (CAD) and type 2 diabetes (T2D). **b,** Per-standard-deviation effect estimates for the disease-optimal PGS across SDoH tertiles. Points denote estimates and error bars indicate 95% confidence intervals.

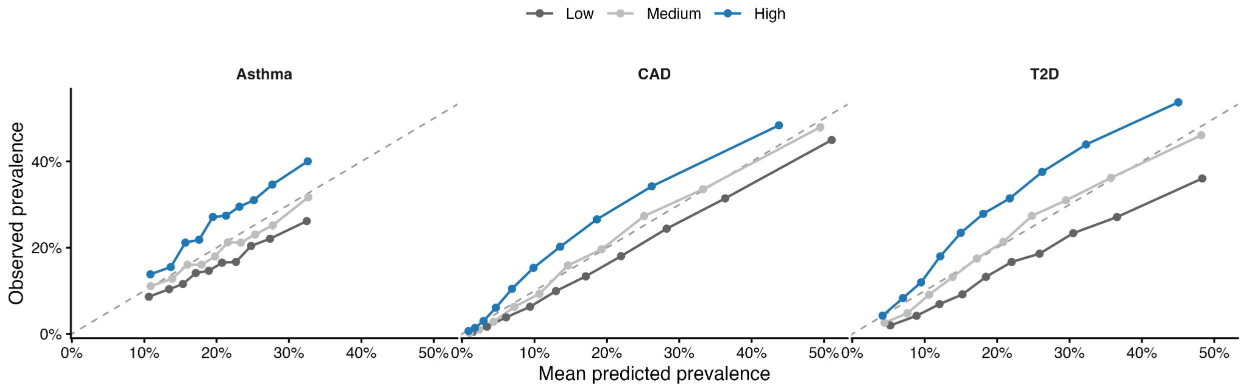


**Extended Data Fig. 11: Calibration of polygenic prediction across social determinants of health strata in AoU-EUR.**

Calibration plots showing the relationship between mean predicted prevalence and observed prevalence across deciles of predicted probability, stratified by tertiles of social determinants of health (SDoH) burden (low, medium, high), for asthma, coronary artery disease (CAD), and type 2 diabetes (T2D). Points represent bin-specific estimates; vertical error bars indicate 95% confidence intervals for observed prevalence, and horizontal error bars indicate 95% confidence intervals for mean predicted prevalence. The dashed line indicates perfect calibration.
