## Supplementary Notes and Figures for "Phenome-derived polygenic scores and social determinants jointly shape context-dependent disease risk"

#### Supplementary Note 1: Enrichment statistics

To characterize the phenomic structure of associations captured by factor-based PGS, we performed enrichment analyses across PheCode chapters. Enrichment was evaluated within each biobank-by-ancestry stratum based on the results of PGS-PheCode association testing. First, within each stratum, we identified PheCodes significantly associated with each factor-based PGS after Bonferroni correction for the total number of tested PheCodes in that stratum. PheCodes were grouped into clinically defined chapters according to the standard PheCode hierarchy. For each factor and PheCode chapter, we assessed whether significant associations were overrepresented relative to expectation given the total number of tested outcomes.

Specifically, for a given factor $f$ and PheCode chapter $c$, we defined $N_f$ as the total number of PheCodes tested for factor $f$, $K_f$ as the number of PheCodes significantly associated with factor $f$, $M_c$ as the number of PheCodes belonging to chapter $c$, and $O_{fc}$ as the observed number of significant associations between factor $f$ and PheCodes in chapter $c$. Under a null model of random distribution of significant associations across PheCodes, the expected number of associations in chapter $c$ is given by $E_{fc} = K_f \times M_c \,/\, N_f$, with variance $Var_{fc} = K_f \times (M_c/N_f) \times (1 - M_c/N_f)$. Enrichment Z-scores were then computed as: $Z_{fc} = \frac{O_{fc} - E_{fc}}{\sqrt{Var_{fc}}}$ , where positive Z-scores indicate overrepresentation of significant associations within a PheCode chapter, and negative Z-scores indicate relative depletion.

To summarize enrichment patterns across biobanks within each ancestry group, Z-scores were meta-analyzed using Stouffer's method[1], weighting each biobank by the square root of the number of PheCodes in the corresponding chapter. Meta-analytic Z-scores were capped at ±6 for visualization. For the main enrichment heatmap, factor-chapter pairs were required to be supported by significant associations in at least two biobanks in European ancestry analyses to emphasize reproducibility; due to more limited representation of non-European ancestry groups across biobanks, enrichment patterns for other ancestries were visualized when supported by at least one biobank.

Enrichment analyses were performed separately for each ancestry group. No additional multiple-testing correction was applied to enrichment Z-scores, as they were used to summarize and visualize phenomic structure rather than to perform formal hypothesis testing.

#### Supplementary Note 2: Variable coding and harmonization

For primary analysis within European ancestry, we selected 29 SDoH variables with low missingness to ensure adequate power for stratified analyses while maintaining comprehensive coverage across all five HP2030 domains (**Table S6**). Variable selection prioritized measures demonstrating monotonic SDoH burden (higher values indicating greater social adversity), conceptual alignment with HP2030 objectives, and adequate response rates.

To address concerns that SDoH associations might reflect downstream consequences of disease (reverse causation), we identified 16 temporally-robust SDoH variables selected for minimal susceptibility to acute disease-related changes  (**Table S6**). These variables were chosen to represent exposures typically established before disease onset or resistant to short-term health shocks. The 16-variable temporally-robust analysis was conducted in European ancestry only.

For cross-ancestry analysis, we employed a reduced 7-variable subset from The Basics survey with <1% missingness across all populations, due to substantially higher missingness rates in African and Admixed American ancestries. This subset provided incomplete domain coverage–omitting social/community context entirely and capturing only 1-2 items from other domains–but ensured adequate sample sizes for ancestry-stratified analyses. Importantly, the 7-variable cross-ancestry analysis used complete case analysis (participants with missing data on any of the 7 items were excluded, representing <1% loss per ancestry) rather than multiple imputation, given the minimal missingness rates for these specific variables.

SDoH variables were recoded to ensure consistent directionality, with higher values uniformly indicating greater social adversity or disadvantage. Binary variables (e.g., lack of health insurance, food insecurity, delayed care due to cost) were coded as 0 (absent/no disadvantage) or 1 (present/disadvantage). Ordinal variables with natural hierarchies were assigned sequential numeric values preserving rank order. Frequency scales (e.g., "never" to "always" for social contact) were coded and oriented so higher values indicate greater social isolation or disadvantage.

For categorical variables representing underlying continuous constructs (e.g., employment status: employed full-time, part-time, unemployed, disabled, retired), we converted responses to ordinal scales reflecting increasing SDoH burden based on economic security and health implications (employed full-time=1, part-time=2, unemployed=3, disabled=4). Housing and neighborhood quality variables were similarly operationalized from categorical descriptors to ordinal burden scales.

Responses indicating "Prefer not to answer," "Skip," or "Don't know" were treated as missing (NA). For the primary 29-variable European ancestry analysis and the 16-variable temporally-robust sensitivity analysis, missing values were addressed via multiple imputation (see below). For the 7-variable cross-ancestry analysis, participants with any missing responses on the seven items were excluded (complete case analysis).

#### Supplementary Note 3: Multiple imputation framework (29-variable and 16-variable analyses)

Missing data in the 29-variable SDoH set and 16-variable temporally-robust set within European ancestry were addressed using multiple imputation by chained equations (MICE) implemented in the mice package (v3.16.0) in R[2]. We generated M=50 imputed datasets to achieve stable parameter estimates and appropriate coverage of confidence intervals, following standard recommendations for multiple imputation.

**Imputation model specification:** Imputation models employed fully conditional specification (FCS), where each variable with missing data is imputed conditional on all other variables in the imputation model. Predictor sets for each imputed variable included: (1) all other SDoH items (ensuring internal consistency within the SDoH construct); (2) all PGS (factor and disease-specific for each anchor outcome) to maintain genetic-social correlations; (3) demographic covariates (age, sex); (4) genetic ancestry (first 10 ancestry-informative principal components); and (5) area-level socioeconomic indicators (median income, poverty, no health insurance, deprivation index, high school education, vacant housing) derived from linked geocoded data to leverage neighborhood-level information for individual-level imputation.

**Imputation methods:** Methods were tailored to variable type to ensure plausible imputations. Predictive mean matching (PMM) was used for continuous variables and ordinal variables with ≥5 categories (e.g., income brackets, education levels), which preserves observed value distributions and avoids out-of-range imputations by selecting imputed values from observed cases with similar predicted values. Logistic regression was used for binary variables (e.g., insurance coverage, food insecurity). Polytomous regression was used for unordered categorical variables with <5 categories (e.g., employment status categories). All continuous auxiliary variables (age, PCs, area-level SES) were mean-centered and scaled to improve imputation model stability.

**Convergence and quality assessment:** Imputation convergence was assessed by visually inspecting trace plots of imputed values across iterations (30 iterations per imputation were used to ensure convergence). All chains showed stable mixing without systematic trends or divergence. Imputation quality was evaluated by comparing distributions of observed versus imputed values for each variable using density plots and summary statistics (mean, SD, quantiles), confirming that imputations were plausible and did not introduce systematic biases.

The 16-variable temporally-robust sensitivity analysis used the same imputation framework but restricted to the 16 specified variables plus all auxiliary predictors. The 7-variable cross-ancestry analysis did not use imputation; instead, participants with any missing data on the seven items were excluded (complete case analysis), which resulted in <1% sample loss per ancestry given the minimal missingness rates for these items.

#### Supplementary Note 4: Control-based re-standardization for predictive modeling

Within each imputation or in the complete dataset, SDoH variables were standardized to ensure equal weighting regardless of original measurement scales and to facilitate comparability across variables with different distributions. Standardization was performed separately within each ancestry group to account for population-specific SDoH distributions and avoid imposing European-centric reference values on AMR and AFR.

To ensure that SDoH effect estimates represent deviations from the disease-free reference population, SDoH composite scores were re-standardized within each imputation and ancestry using the mean and standard deviation from control individuals (disease-negative for the anchor outcome being analyzed): $SDoH_Z = (SDoH_{composite} - \mu_{controls}) / \sigma_{controls}$. This control-based standardization parallels the approach used for PGS standardization and ensures that reported effect sizes (log-odds, odds ratios) represent the increase in disease odds per standard deviation of SDoH burden observed in the unaffected reference population.

The re-standardized $SDoH_Z$ scores were used in all logistic regression models, interaction analyses, and stratification procedures. For imputed analyses (29-variable and 16-variable), models were fit separately within each of the M=50 imputations and results were pooled using Rubin's rules. For the 7-variable cross-ancestry analysis using complete cases, models were fit once on the complete dataset.

#### Supplementary Note 5: Composite score validation

To quantify information loss from reduced SDoH representations, we benchmarked the 7-variable and 16-variable composites against the full 29-variable composite in European ancestry. Benchmarking was conducted by comparing variance explained when each composite was used to predict disease status in covariate-adjusted logistic regression models (adjusted for age, sex, and 10 genetic PCs). The 7-variable composite retained 58-86% of the predictive variance captured by the 29-variable composite across diseases (asthma: 58%; CAD: 86%; T2D: 96%). The 16-variable temporally-robust composite retained 80-100% of the predictive variance across diseases (asthma: 80%; CAD: 95%; T2D: 100%), confirming that temporally-robust variables captured the majority of SDoH-disease associations while minimizing reverse causation concerns.

These benchmarking results indicate that: (1) cross-ancestry analyses using the 7-variable composite provide conservative estimates of SDoH contribution, likely underestimating the true effect due to incomplete domain coverage; and (2) sensitivity analyses using temporally-robust variables preserve most of the predictive signal while addressing reverse causation, supporting the validity of observed SDoH effects as antecedent rather than consequent to disease.

#### Supplementary Note 6: Gene-Environment Interaction Analysis

To evaluate whether SDoH modifies the realized expression of genetic liability, we tested for multiplicative interaction between disease-optimal PGS and SDoH composite using logistic regression. For each disease and ancestry, we fit the following interaction model within each imputation:

$$Logit(P\ (Disease\ =\ 1))\ = \beta_0\ + \beta_{PGS}PGS\ + \beta_{SDoH}SDoH\ + \beta_{int}(PGS\ \times\ SDoH)\ + C\ ;$$

where PGS and SDoH were standardized to mean 0 and SD 1, and $C$ represents covariates. The interaction coefficient $\beta_{int}$ quantifies the change in the log-odds of the PGS effect per SD increase in SDoH burden. Significance of interaction was assessed using Wald test of $\beta_{int}$. Negative interaction coefficients indicate attenuation of genetic effects under higher social adversity; positive coefficients indicate amplification. ORs and 95% confidence intervals were computed by exponentiating the pooled log-odds coefficients and their confidence bounds. Because both PGS and SDoH were standardized, reported ORs represent the multiplicative change in disease odds per one standard deviation increase in each predictor.

**Age- and sex-stratified interaction analyses.** We repeated the PGS×SDoH interaction models within prespecified age (<50, 50-65 and >65 years) and sex strata. Within each subgroup, we refit logistic regression models including standardized SDoH, standardized PGS and their multiplicative interaction term, adjusting for the same non-stratifying covariates and ancestry principal components as in the primary analyses. PGS were standardized using the control distribution, SDoH was re-standardized within each imputation using controls only, and analyses were restricted to the shared set of participants with complete phenotype, PGS, covariate and principal component data and non-missing SDoH across all imputations. Covariate terms derived from the stratifying variable were removed before model fitting, and interaction coefficients were pooled across imputations using Rubin’s rules.

#### Supplementary Note 7: Stratified Performance Analysis by SDoH Burden

##### SDOH tertile construction

To evaluate disease risk across joint strata of genetic and social burden, we partitioned participants into tertiles of SDoH burden within each ancestry. For the primary European ancestry analysis using 29 SDoH variables and 16-variable analysis, tertile boundaries were defined using a quantile-based stratification scheme: within each imputation, individuals were ranked by their SDoH composite score and assigned to tertiles (bins 1, 2, 3) based on empirical 33rd and 67th percentiles, ensuring approximately equal sample sizes per stratum. This per-imputation stratification accounts for the variability in SDoH distributions across imputed datasets while maintaining consistent relative rankings.

PGS was independently binned into deciles (10 equal-frequency bins) within each ancestry and imputation using the same quantile-based approach, yielding a 10×3 joint stratification grid (PGS decile × SDoH tertile) for standardized predicted prevalence surface analysis.

##### Stratified discrimination and PGS effect attenuation

Within each SDoH tertile,we evaluated the incremental discrimination provided by adding PGS to the baseline covariate model. We fitted two logistic regression models: (i) BASE model including covariates only; 2) BASE+optimal PGS model. Incremental predictive performance (*Δ*AUC) was calculated as the difference between the PGS model and baseline model.

To quantify genetic stratification on the odds-ratio scale, we compared disease prevalence between individuals in the top PGS decile (decile 10) versus bottom PGS decile (decile 1) within each SDOH tertile. Odds ratios and 95% confidence intervals were computed via logistic regression (Disease ~ $PGS_{decile}$ + covariates) within each tertile and imputation, with coefficients pooled across imputations using Rubin's rules. We additionally calculated per-standard-deviation ORs ($\beta_{PGS}$ from logistic regression with continuous $PGS_Z$) to assess linear genetic effects across the full PGS distribution.

**Attenuation metrics:** The percentage change in $\Delta$AUC and OR from the lowest to highest SDOH tertile quantifies effect modification. For example, $[(\Delta AUC_{low} - \Delta AUC_{high})/\Delta AUC_{low}] \times 100\%$ represents the percent attenuation in genetic discrimination under high versus low social adversity.

#### Supplementary Note 8: Standardized predicted-prevalence grids and joint stratification

To visualize disease prevalence across the joint genetic-social space, we estimated observed prevalence and standardized predicted prevalence within each cell of a 10 × 3 grid defined by PGS decile and SDoH tertile. Within each imputation, observed prevalence in each cell was calculated as the proportion of cases among individuals in that cell:

$$prev_{obs} = \frac{n_{cases}}{n_{total}}.$$

Standardized predicted prevalence was obtained by fitting the following joint stratification model:

$$\text{Disease} \sim \text{factor(PGS decile)} \times \text{factor(SDoH tertile)} + \text{covariates}.$$

This multiplicative interaction model allows the effect of genetic burden to vary across SDoH strata. To reduce sensitivity to differences in covariate composition across genetic-social strata, predicted values for each cell were obtained by marginal standardization over a common covariate distribution rather than by using the raw observed prevalence in each cell. Specifically, fitted probabilities were generated for all individuals under each PGS decile × SDoH tertile combination while holding all other covariates, including age, at their observed values; age was modeled using the same specification as in the primary outcome models. Cell-level standardized predicted prevalence was then calculated as the mean of these fitted probabilities. This yields a covariate-standardized estimate of disease prevalence for each grid cell, isolating the joint contributions of genetic and social stratification from differences in covariate composition.

For comparison, we also fit an additive model without the interaction term:

$$\text{Disease} \sim \text{factor(PGS decile)} + \text{factor(SDoH tertile)} + \text{covariates},$$

to evaluate whether inclusion of the interaction term improved the fit of the joint prevalence surface.

All standardized predicted prevalences were calculated separately within each imputation and pooled across imputations by taking the mean as the point estimate and the empirical 2.5th and 97.5th percentiles as the 95% confidence interval for each grid cell.

**Predicted prevalence difference.** To quantify genetic stratification within each SDoH tertile, we calculated the difference in standardized predicted prevalence between the highest and lowest PGS deciles:

$$\Delta prev = \hat{P}_{std}(decile\ 10) - \hat{P}_{std}(decile\ 1).$$

This metric summarizes the separation in predicted prevalence between high- and low-genetic-risk groups within a given social context. Predicted prevalence differences were calculated separately within each imputation using the standardized predicted prevalences described above and pooled across imputations using the mean and empirical 95% confidence interval. Larger positive values indicate stronger genetic stratification on the predicted-prevalence scale, whereas attenuation of this difference indicates reduced separation between high- and low-genetic-risk groups within a given SDoH stratum.

#### Supplementary Note 9: Calibration assessment across SDoH strata

To assess whether joint PGS + SDoH models were well calibrated across social contexts, we evaluated calibration separately within each SDoH tertile. Within each imputation, we fit the joint model

$$\text{Disease} \sim \text{PGS}_Z + \text{SDoH}_Z + \text{covariates}$$

and obtained fitted probabilities for all individuals. Calibration was then assessed within each SDoH tertile by comparing fitted probabilities with observed outcomes. We summarized calibration using the calibration slope, calibration intercept, and Brier score. Calibration metrics were computed independently within each imputation and pooled by taking the mean and empirical 95% confidence interval (2.5th-97.5th percentiles) across the (M=50) imputations.

Differences in calibration intercept across SDoH tertiles indicate whether a model trained in the full analysis sample systematically over- or underestimates disease prevalence in different social contexts. For example, negative intercepts in lower-SDoH strata together with positive intercepts in higher-SDoH strata indicate relative overprediction in socially advantaged groups and underprediction in socially disadvantaged groups. Because the main displays of joint genetic-social effects are based on covariate-standardized predicted prevalence, these calibration summaries reflect socially contingent shifts in baseline prevalence beyond simple differences in age composition or other covariates across strata.

Calibration plots were constructed by binning individuals according to predicted probability within each SDoH tertile and comparing mean predicted prevalence with observed prevalence in each bin. The dashed identity line (y=x) indicates perfect calibration.

#### Supplementary Note 10: Tail-prevalence analysis and high-risk group identification

To evaluate the ability of PGS and joint PGS + SDoH models to identify individuals with elevated disease burden across SDoH contexts, we examined the upper tails of the PGS and model-predicted distributions within each SDoH tertile and imputation. Tail groups were defined using three thresholds: the top 10% (90th percentile), top 5% (95th percentile), and top 1% (99th percentile).

Tail groups were defined as follows:

- **Predicted-prevalence tail:** individuals in the top (X%) of fitted probabilities from the joint PGS + SDoH + covariates model
- **PGS tail:** individuals in the top (X%) of the disease-optimal PGS distribution, irrespective of SDoH

For each tail group, we calculated observed prevalence and compared it with the baseline prevalence within the corresponding SDoH tertile to quantify prevalence enrichment. A minimum sample size of (n=200) per tertile was required to compute tail summaries, to ensure stable prevalence estimates.

The following tail metrics were calculated within each SDoH tertile:

1. **Observed prevalence in tail:** the proportion of cases among individuals in the tail group
2. **Prevalence enrichment:** the ratio of tail prevalence to the baseline prevalence in the same SDoH tertile
$$Enrichment = \frac{tail\ prevalence}{tertile\ baseline\ prevalence}$$
Values greater than 1 indicate that the tail identifies individuals with higher disease prevalence than the average within that social stratum.
3. **Mean score in tail:** the mean fitted probability or mean PGS value within the tail group, used to confirm appropriate tail identification

Tail metrics were calculated separately within each imputation and pooled using the mean and empirical 95% confidence interval. Comparing prevalence enrichment across SDoH tertiles allowed us to assess whether high genetic burden retained its predictive value under social adversity or whether this value was attenuated in higher-SDoH contexts.

#### Supplementary Note 11: Incident-case Cox proportional hazards analyses

As a prospective sensitivity analysis, we fit incident-case Cox proportional hazards models in All of Us for asthma, CAD, and T2D. Individuals with prevalent disease at baseline were excluded. Follow-up time was defined from enrollment to incident diagnosis, death, last follow-up, or administrative censoring.

Models adjusted for baseline age, age squared, sex, age-by-sex, age-squared-by-sex, and the top 10 genetic principal components. Factor-derived and disease-specific PGS, as well as the SDoH composite, were standardized within ancestry prior to modeling. Nested Cox models evaluated baseline covariates alone; covariates plus each PGS separately; covariates plus SDoH; additive joint models including PGS and SDoH; and interaction models including PGS×SDoH terms. Joint models containing both PGS representations were also evaluated.

Hazard ratios and 95% confidence intervals were reported per standard deviation increase in predictor values. Prospective discrimination was summarized using Harrell's concordance index and time-dependent AUC at prespecified follow-up horizons. For analyses involving multiple imputation, models were fit separately within each imputed dataset and summary estimates were combined across imputations. These analyses were intended to assess whether the joint genetic-social risk architecture observed in the primary case-control analyses extended to future disease risk.

### Supplementary Figures

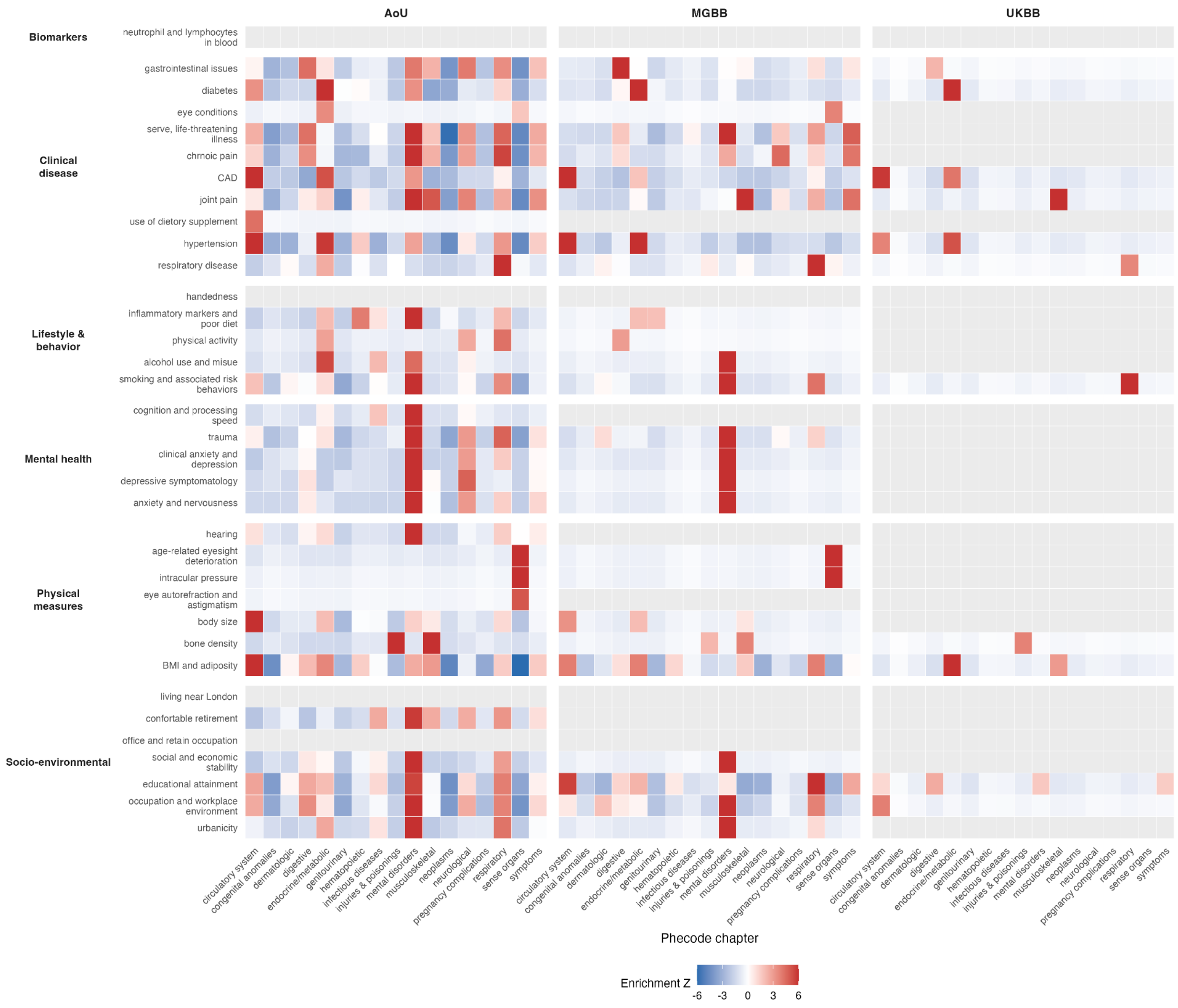


**Figure S1. Phenomic enrichment of factor-based polygenic scores across biobanks in European ancestry populations.**

Heatmaps show enrichment Z-scores summarizing the concentration of significant associations between factor-based PGS and phecode chapters in individuals of European ancestry, shown separately for the UK Biobank (UKB) holdout, the Mass General Brigham Biobank (MGBB), and the All of Us Research Program (AoU). Rows correspond to the 35 latent phenomic factors grouped into six broad domains, and columns correspond to phecode chapters. Enrichment Z-scores were computed within each biobank based on Bonferroni-significant PGS-phecode associations using a hypergeometric framework (**Supplementary Note 1**). Color indicates the direction and magnitude of enrichment, with positive values reflecting overrepresentation of associations within a chapter and negative values indicating relative depletion. Z-scores were capped at ±6 for visualization.

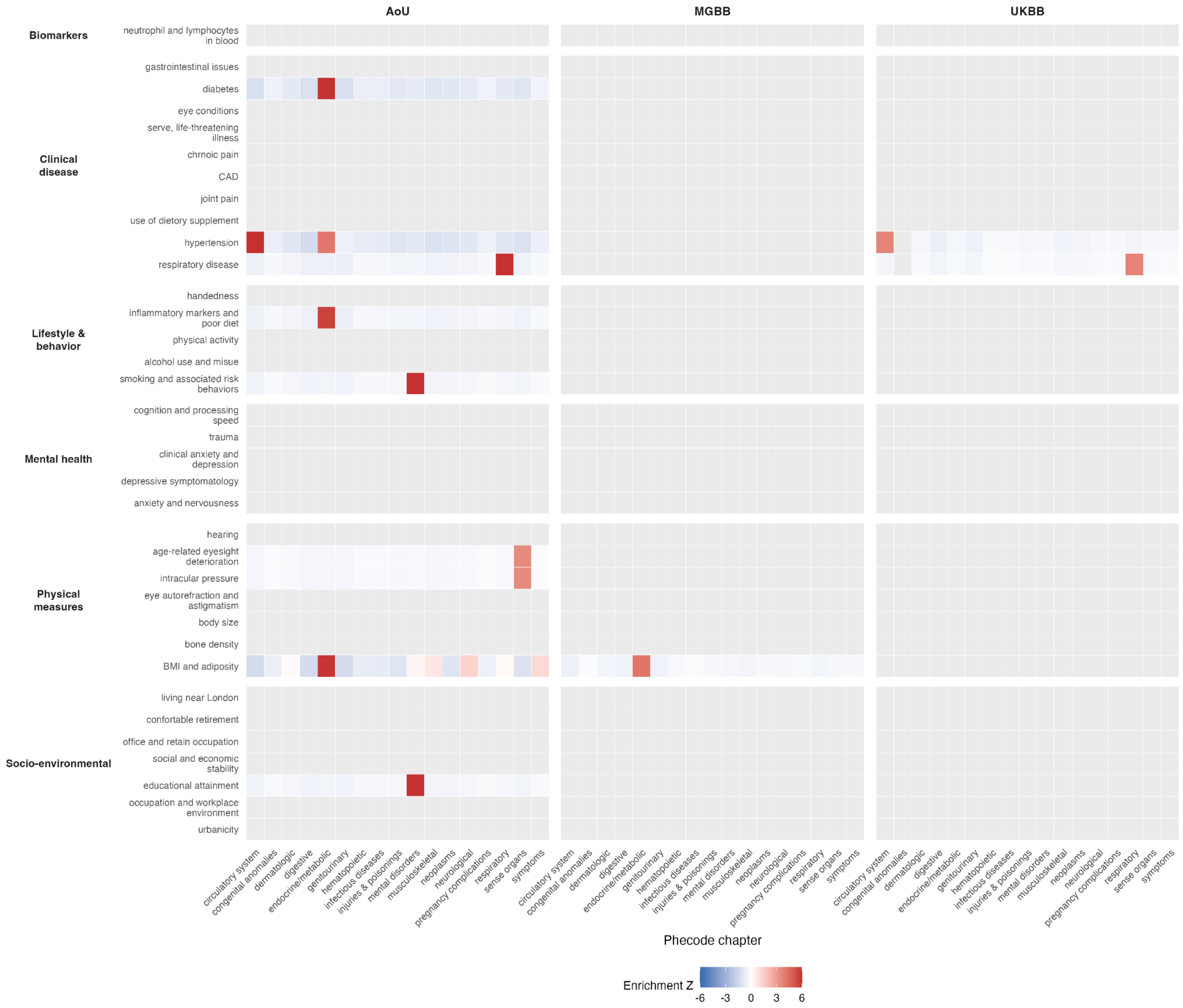


**Figure S2. Phenomic enrichment of factor-based polygenic scores across biobanks in African ancestry populations.**

Heatmaps show enrichment Z-scores summarizing the concentration of significant associations between factor-based PGS and phecode chapters in individuals of African ancestry, shown separately for the UK Biobank (UKB) holdout, the Mass General Brigham Biobank (MGBB), and the All of Us Research Program (AoU). Rows correspond to the 35 latent phenomic factors grouped into six broad domains, and columns correspond to phecode chapters. Enrichment Z-scores were computed within each biobank based on Bonferroni-significant PGS-phecode associations using a hypergeometric framework (**Supplementary Note 1**). Color indicates the direction and magnitude of enrichment, with positive values reflecting overrepresentation of associations within a chapter and negative values indicating relative depletion. Z-scores were capped at ±6 for visualization.

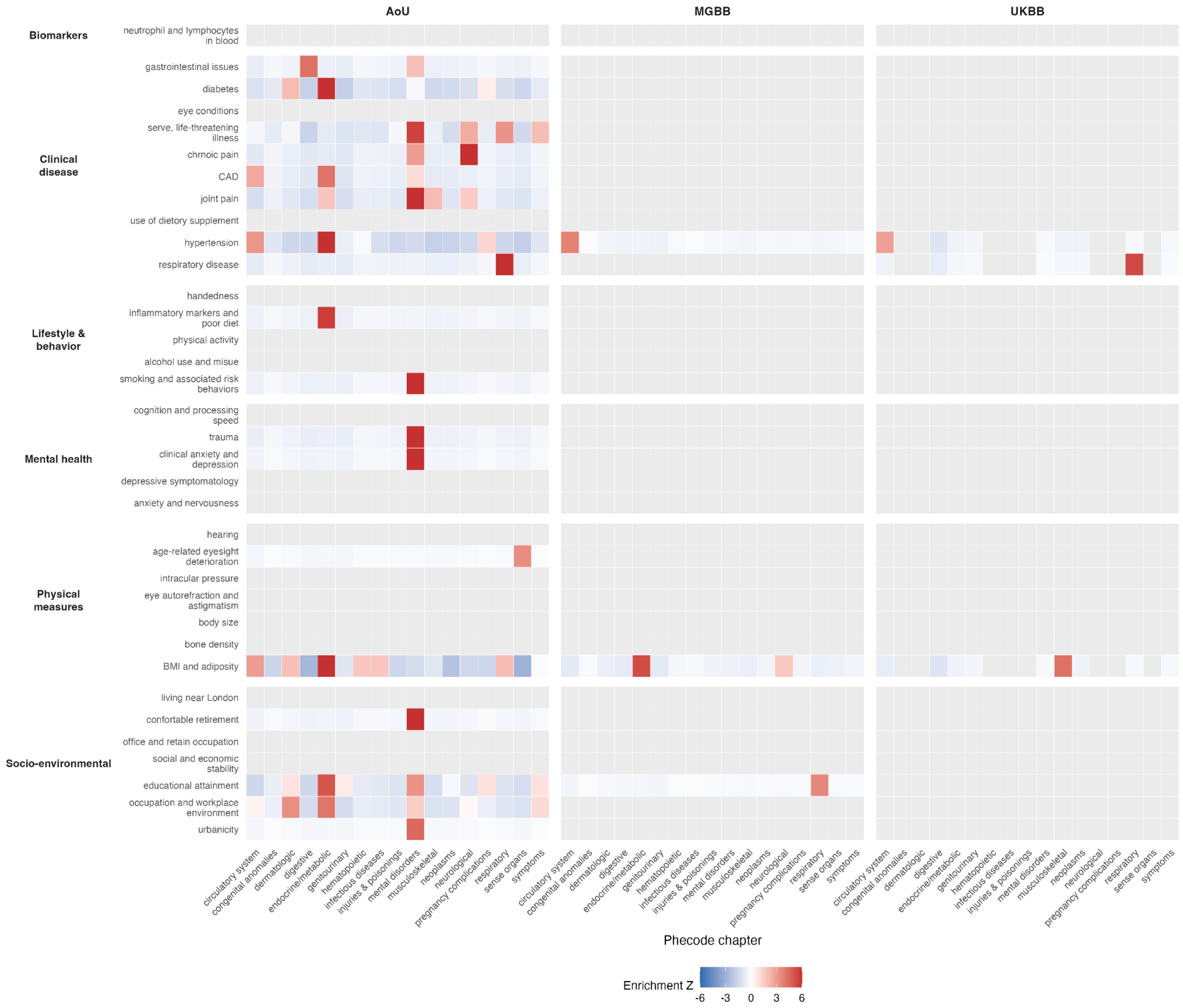


**Figure S3. Phenomic enrichment of factor-based polygenic scores across biobanks in Admixed American ancestry populations.**

Heatmaps show enrichment Z-scores summarizing the concentration of significant associations between factor-based PGS and phecode chapters in individuals of admixed american ancestry, shown separately for the UK Biobank (UKB) holdout, the Mass General Brigham Biobank (MGBB), and the All of Us Research Program (AoU). Rows correspond to the 35 latent phenomic factors grouped into six broad domains, and columns correspond to phecode chapters. Enrichment Z-scores were computed within each biobank based on Bonferroni-significant PGS-phecode associations using a hypergeometric framework (**Supplementary Note 1**). Color indicates the direction and magnitude of enrichment, with positive values reflecting overrepresentation of associations within a chapter and negative values indicating relative depletion. Z-scores were capped at ±6 for visualization.

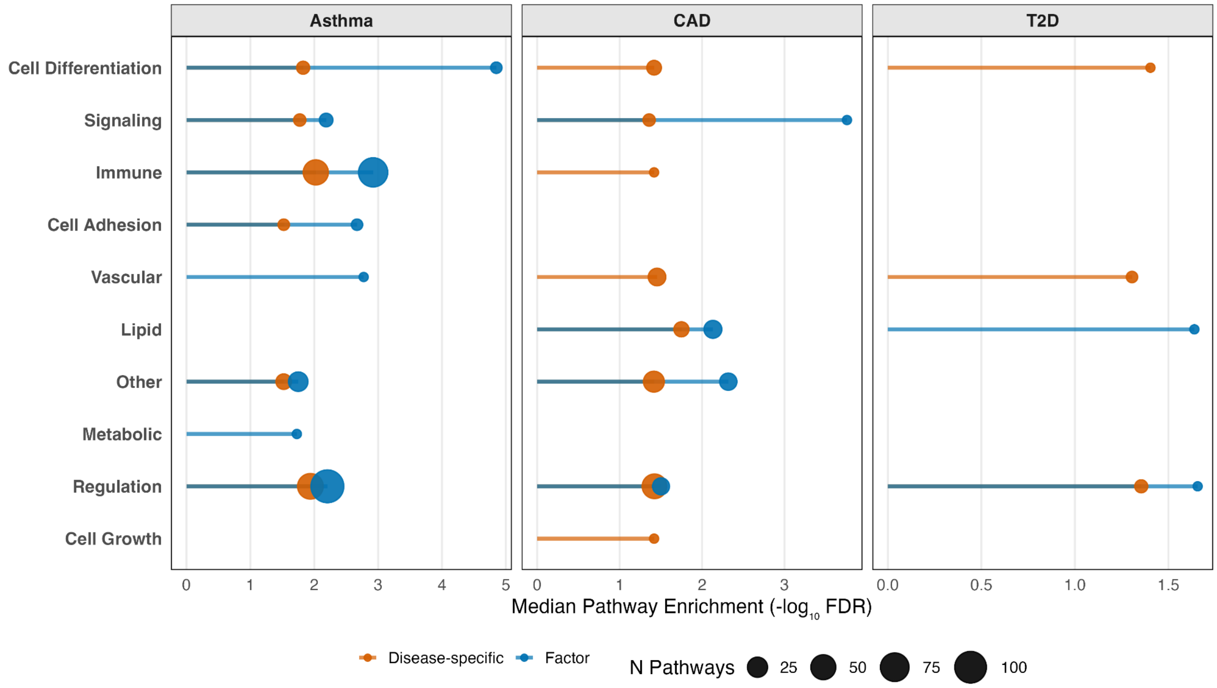


**Figure S4. Functional category profiles of MAGMA pathway enrichments for factor-versus disease-specific GWAS.**

MAGMA gene-set enrichment results were grouped into broad biological categories based on Gene Ontology pathway names (y-axis). For each trait (asthma, coronary artery disease (CAD), and type 2 diabetes (T2D); facets), points show the median enrichment strength within a category, summarized as median -$\log_{10}$FDR across all pathways in that category, for the factor-based GWAS (blue) and disease-specific GWAS (orange). Point size is proportional to the number of pathways contributing to the category.

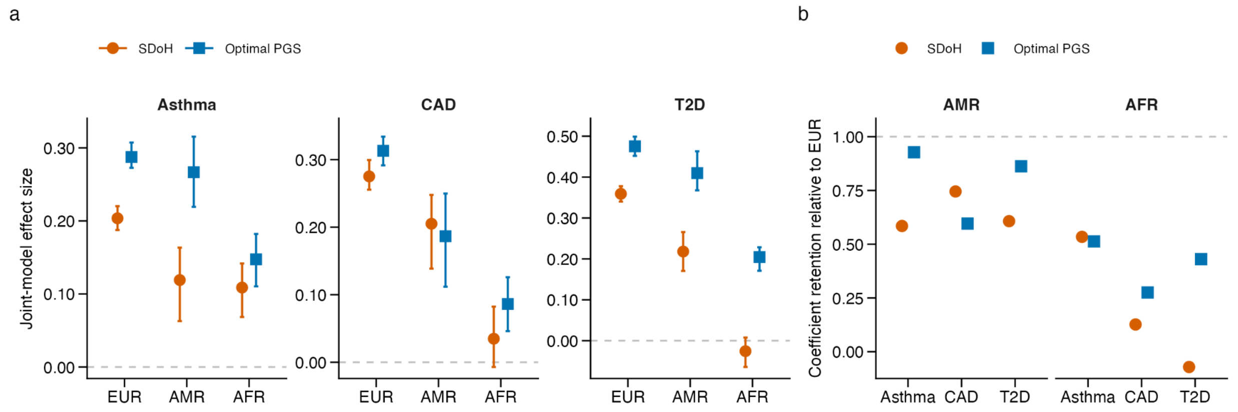


**Figure S5. Cross-ancestry comparison of joint-model effect estimates under a harmonized SDoH composite.**

**a,** Joint-model coefficients for the harmonized 7-variable social determinants of health (SDoH) composite and the ancestry-specific optimal polygenic score (PGS) across European (EUR), Admixed American (AMR) and African (AFR) populations. Asthma models use the factor-based PGS, whereas coronary artery disease (CAD) and type 2 diabetes (T2D) use disease-specific PGS. Points denote effect estimates and error bars indicate 95% confidence intervals.

**b,** Retention of joint-model effect estimates relative to European ancestry. Coefficients for SDoH and the optimal PGS are expressed as ratios relative to their corresponding estimates in EUR. Values below 1 indicate attenuation relative to EUR.

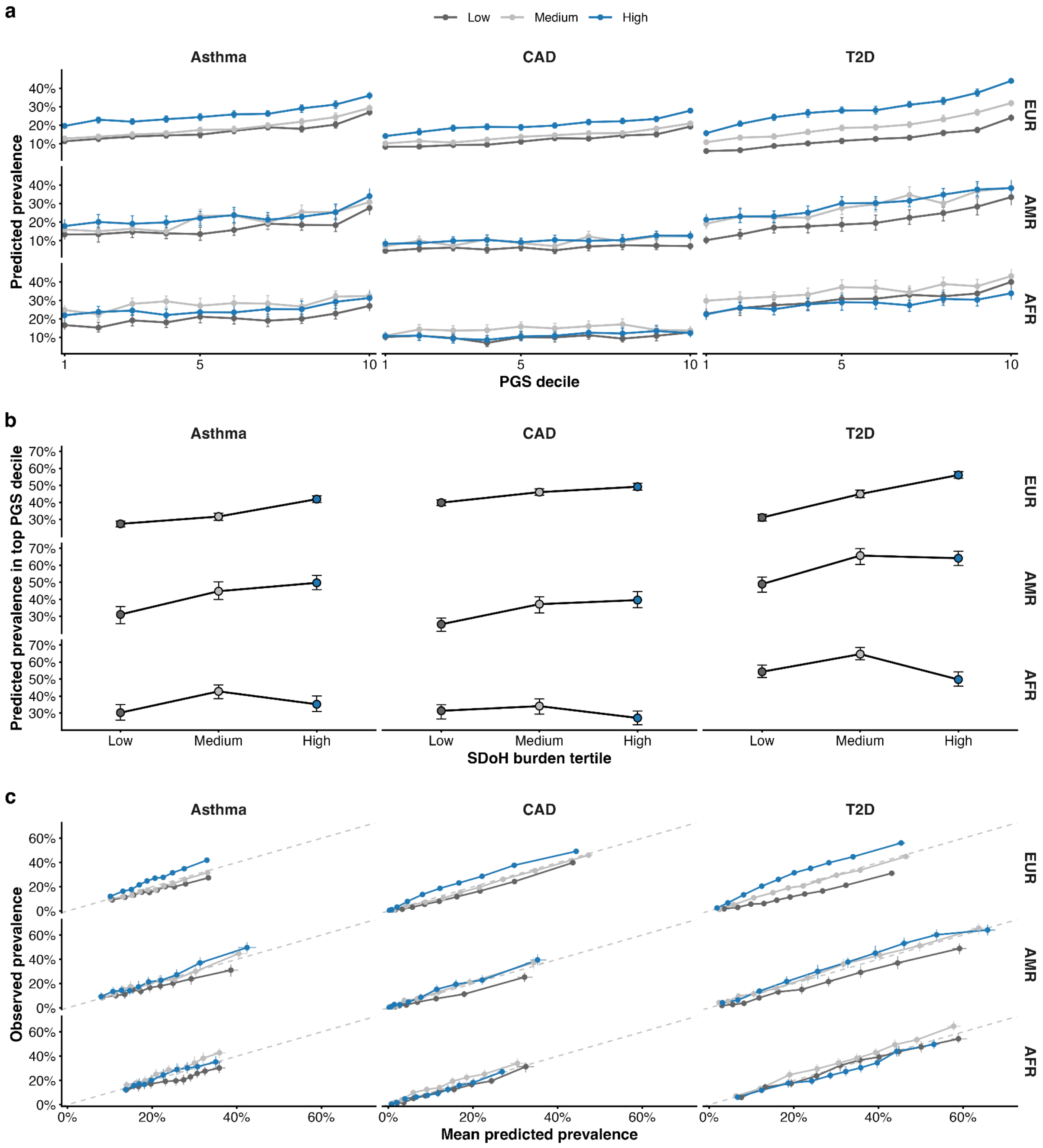


**Figure S6. Cross-ancestry predicted-prevalence and calibration patterns under the harmonized SDoH composite.**

**a,** Standardized predicted prevalence across deciles of the disease-optimal polygenic score (PGS) within tertiles of social determinants of health (SDoH) burden (low, medium, high), stratified by ancestry (European (EUR), Admixed American (AMR), African (AFR)) and disease (asthma, coronary artery disease (CAD), and type 2 diabetes (T2D)). Lines connect decile-specific estimates within each SDoH stratum; points indicate estimates and error bars indicate 95% confidence intervals.

**b,** Standardized predicted prevalence among individuals in the highest PGS decile across SDoH tertiles, stratified by ancestry and disease. Points indicate estimates and error bars indicate 95% confidence intervals.
**c,** Calibration plots showing observed prevalence versus mean predicted prevalence across calibration bins within each SDoH tertile, stratified by ancestry and disease. The dashed line indicates perfect calibration (y=x). Points indicate bin-level estimates and error bars indicate 95% confidence intervals.

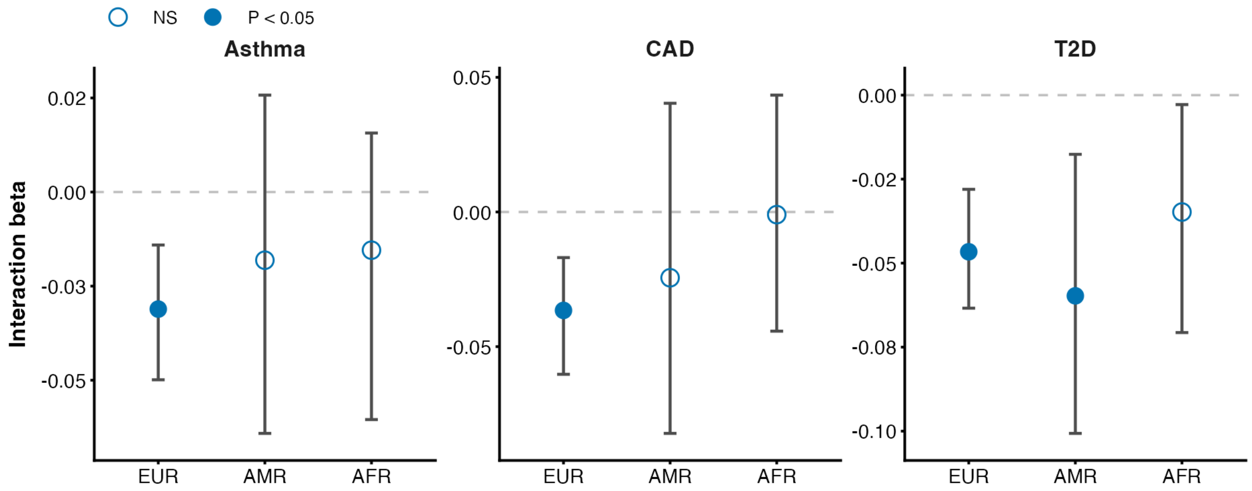


**Figure S7. Cross-ancestry interaction estimates under the harmonized SDoH composite.** Interaction coefficients and 95% confidence intervals for the product term between the harmonized SDoH composite and the ancestry-specific optimal PGS across European (EUR), Admixed American (AMR) and African (AFR) populations. Asthma uses the factor-based PGS; CAD and T2D use disease-specific PGS. Filled points indicate $P < 0.05$.

### References


1. Stouffer, S. A. A study of attitudes. *Sci. Am.* **180**, 11–15 (1949).
2. van Buuren, S. & Groothuis-Oudshoorn, K. mice: Multivariate Imputation by Chained Equations inR. *J. Stat. Softw.* **45**, (2011).
